# The covariance matrix of metapopulation disease models and applications to early warning signals

**DOI:** 10.64898/2026.05.08.26352721

**Authors:** Joshua Looker, Kat S. Rock, Louise Dyson

## Abstract

Infectious disease time series often show signs of epidemic transitions, such as the peaks and troughs of the time series. In these time series, key system parameters can lead to catastrophic changes in the dynamical system behaviour (often called critical transitions). Modellers have increasingly shown that early warning signals can anticipate these transitions, both critical and non-critical, in infectious disease time series. Existing methods, however, generally focus on univariate time series data, or ignore spatiotemporal patterns that may be present as a disease spreads through a population. Recent ecological literature developments expand existing temporal and spatial methods to consider the covariance matrix of multiple, related time series. However, many of these proposed signals still make an assumption of stationary time series/system equilibrium. Whilst often true in ecological modelling, disease systems are seldom at equilibrium. In this paper, we propose the usage of the eigendecomposition of the non-stationary covariance matrix as a more suitable early warning signal for epidemiological data. We first analyse the expected trends in the eigenvalues and eigenbasis of the covariance matrix on approach to a transition. Next we apply these methods to a spatially-structured susceptible-infectious-recovered model to explore how the eigenbasis may provide extra information to modellers. Finally, we test these methods on SARS-CoV-2 case data during the 2020–2021 pandemic period in England.

## Introduction

Epidemiological time series often display periods of unpredictable behaviour, possibly caused by intervention measures or the introductions of new viral strains^1,2^. Of particular interest to modellers is considering whether the disease is approaching elimination or (re-)emergence. These so-called critical transitions are often functions of an underlying, unobservable parameter in the system^3^. As the parameters change, so does the stability of system equilibria (for epidemiology, these correspond to the stability of a disease-free and endemic equilibria), leading to a bifurcation. The stability of these equilibria is found from the real part of the eigenvalues of the Jacobian and are direct functions of the system parameters. Of particular interest to infectious disease modellers is the basic reproduction number (*R*_0_). When this is greater than one, the endemic equilibrium is stable and the disease may persist in the population. However, when the basic reproduction number is less than one, the disease-free equilibrium is stable and the disease may die out. Thus, *R*_0_ = 1 represents a bifurcation in the system where a critical transition from disease persistence to disease elimination may occur^2^. Non-pharmaceutical interventions (NPIs), vaccinations, and other measures may be taken by governing bodies to help control the spread of the pathogen, possibly changing *R*_0_ and leading to a bifurcation. Although transitions may happen quickly, they are often preceded by predictable behaviour in statistics of the observed cases, which can help in anticipating bifurcations and is an early warning signal (EWS) of an upcoming critical transition. Additionally, EWSs have been shown to also anticipate non-critical transitions^4–9^. In infectious disease modelling, these non-critical epidemic transitions correspond to whether the epidemic is growing or declining, represented by whether the effective reproduction number (*R*_*t*_) is greater than or less than one. Indeed, we have previously shown the ability of EWSs to anticipate these epidemic transitions, such as peaks in infection curves^4^.

Many such EWSs are based on the theory of critical slowing down. This predicts that system oscillations take longer to return to the long-term equilibrium on the approach to a critical transition, leading to signals such as an increase in the variance of the system state. Early warning signals based on multivariate time series have also been developed and applied to various modelling areas^10–14^. Some of these generalise the increasing variance of a univariate time series by focusing on the eigenvalues and eigenvectors of the covariance matrix of interlinked time series, such as the fish population in different sub-locations of a reservoir. The modelling predicts monotonic changes in the eigenvalues on approach to a critical transition and it is suggested that the eigenvectors, and signals calculated from the corresponding primary component analysis (PCA), may also provide extra information to modellers in identifying at risk sub-populations that have the most impact on potential critical transitions^10,13,15–18^. In infectious disease modelling, it is common to aggregate/disaggregate case data to account for pathogen spread through different sub-populations and to reduce stochasticity. Further, the collection of case data is often focused on certain age-groups or regions, especially when targeting elimination. The recent developments discussed above suggest that having high granularity and disaggregation could be useful in informing covariance matrix-based EWSs.

To our knowledge, the current literature does not consider the effects of non-stationary stochastic oscillations on the covariance matrix. Explicitly, Chen *et al*.^11^ analyse the eigenvalues of the covariance matrix for an ecological system transitioning from its long-term equilibrium towards system collapse. O’Dea *et al*.^19^ examine how the eigendecomposition of the covariance between susceptibles and infectious individuals could anticipate the distance to a critical transition for disease systems with fewer compartments and systems at steady-state disease systems. Both build on the original results by Kwon *et al*.^20^, but all assume that the stochastic fluctuations in the dynamic system have reached a stationary state and the system is oscillating around its stable equilibrium point. This allows for the analytical calculation of the covariance matrix eigenvalues as a function of the bifurcation parameter. However, this implicitly assumes that a timescale separation is possible; such as that often undertaken in ordinary differential equation (ODEs) models with a fast-slow bifurcation system. In many metapopulation disease models and in real disease case data, this may not be possible. For a slower bifurcation (such as when a disease is targeted for elimination), the demographics of the population may be shifting at a similar rate to the bifurcation parameter, and so a timescale separation may not be possible. Further, the timescale separation requires that we know the state the fast system will reach. As the timescales may be similar and the endemic equilibrium is often close to the disease-free equilibrium, noise-induced tipping may occur and the fast system may not behave as expected^21^. Finally, by definition, during (re-)emergence of novel diseases, the stochastic disease system is non-stationary (*i*.*e*. not at equilibrium) and so the usual stationary assumption will not apply.

Building on the current literature, this paper explores the behaviour of a non-stationary covariance matrix of metapopulation-structured disease models on approach to an epidemic transition. We consider the time evolution of the eigendecomposition of the covariance matrix, and show that there are predictable trends in the dominant eigenvalue(s) of the system, even for non-stationary fluctuations. We also propose the rate of rotation of the eigenbasis of the covariance matrix as an additional EWS and show that changes in the dominant eigenvector may provide extra information on which groups contribute most to future infection dynamics. Finally, we analyse the behaviour of these proposed covariance matrix-based EWSs on case data from the 2020–2021 period of the SARS-CoV-2 pandemic in England.

## Results

### Eigendecomposition of the Covariance Matrix

The spread of an infectious pathogen through a population is often modelled by splitting people up into smaller sub-populations. These sub-populations could represent numerous characteristics, but are most often used to separate people by age and location. Metapopulation and age-structured models were used extensively during the SARS-CoV-2 (COVID-19) pandemic to simulate the spread of the disease through different locations. Such compartment models are often represented by a system of ODEs in the form

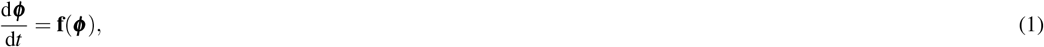

which expresses the deterministic evolution of the state vector ***ϕ*** (*t*) of the system subject to the vector of model equations **f**. However, many real-world examples of these systems display intrinsic stochasticity not captured by these deterministic equations. Let ***x*** = *N* ***ϕ*** + *N*^1/2^ ***ξ*** where ***x*** corresponds to the observed system state (including stochasticity), *N* represents the system size (often the population), ***ϕ*** represents the expected solutions to the deterministic equations and ***ξ*** are the stochastic deviations from these solutions. We can use van Kampen’s system-size expansion^22^ to derive the linear Fokker-Planck equation for the probability density, Π(***ξ***,*t*), of these stochastic deviations:

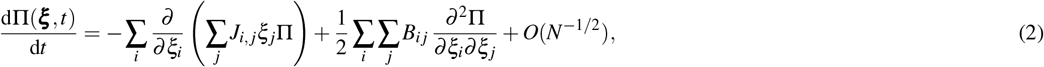

where **J** is the Jacobian of ***f*** and ***B*** is a diffusion matrix derived using the system-size expansion. We also note that **B** and **J** are then independent of ***ξ*** such that ***ξ*** is a mean-zero multivariate Gaussian with a covariance matrix (Σ) satisfying the time-varying Lyapunov equation given by

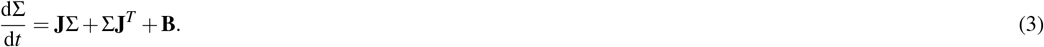

It is here that most existing literature assumes that the fluctuations have reached a stationary distribution; that the system is fluctuating around its long-term equilibrium. This simplifies the above equation such that 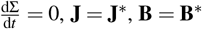 where **J**^∗^ and **B**^∗^ are constant matrices corresponding to the values of the Jacobian and diffusion matrix calculated at equilibrium. However, as previously stated, disease systems going through bifurcation are normally non-stationary especially on the modelling time scales of most disease outbreaks. Our main results consider the more general non-stationary case for the distribution of ***ξ*** . It is worth noting that these results still make the standard assumption that the original system can be approximated by the linear Fokker-Planck equation (both in terms of the detrended and observed time series), which has been shown to be a good approximation for many infectious disease models^4,9,23–27^.

Denoting the eigenvalues of the covariance matrix in descending order as *λ*_1_, …, *λ*_*n*_, then Chen *et al*.,^11^ have shown that *λ*_1_ (or the real parts of the complex conjugate pair *λ*_1_, *λ*_2_ for the case of complex eigenvalues of **J**) will increase on approach to a bifurcation. We build on these results by showing that similar trends can be seen for the non-stationary evolution of Π. We also find that the percentage of the total variation accounted for by the largest eigenvalue may decrease on approach to bifurcation (the reverse of that found by Chen *et al*.). This may be because changes in the eigenmodes of the covariance matrix are necessary to account for the new dynamics introduced by the bifurcation.

For the same reasons, we also propose that rotations in the eigenbasis (for a non-stationary covariance matrix) can also help anticipate upcoming epidemic transitions. These correspond to the evolution of the eigenmodes due to the changing dynamics in the system. In Supplementary Section S4 we show that the dominant eigenvector of the covariance matrix grows exponentially with respect to the projection of the Jacobian onto the covariance eigenbasis, suggesting that changes in the dominant eigenvector are EWSs of a transition.

### Theory: Spatially-structured disease model

To illustrate our proposed EWSs, we begin with a simple model of three interacting metapopulations each undergoing susceptible-infectious-recovered (SIR) disease dynamics with demography. This builds on previous modelling work considered by Rozhnova *et al*.^23,24^ by investigating the covariance matrix on approach to a bifurcation. The model consists of *n* cities, each with a constant population of size *S* _*j*_ + *I*_*j*_ + *R*_*j*_ = *N*_*j*_, *j* = 1, 2,…, *n*, where *S* _*j*_, *I*_*j*_, *R*_*j*_ are the number of susceptible, infectious and recovered individuals in metapopulation *j* respectively. Individuals recover at rate *γ* and are born and die at rate *µ*, where every death event corresponds to a birth event so that the population size remains constant. This model can then be represented by the following set of ODEs (where we note that the *R*_*j*_ equations are omitted as they can be solved for via *N*_*j*_ − *I*_*j*_ − *S* _*j*_):

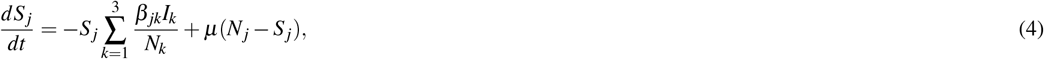

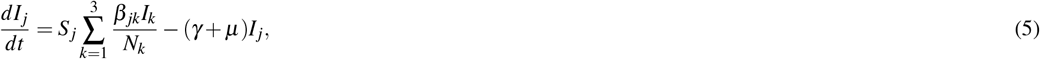

where *β*_*jk*_ is the effective contact rate between susceptibles in population *j* and infectious individuals in population *k*. As a simplification, we set *N*_*j*_ = *N*_*a*_ ∀ *j* such that each metapopulation is of the same size, *N*_*a*_. We further simplify the model by considering a network of three cities on a line (such that transmission occurs only between adjacent cities). We now take the system-size expansion by making the ansatz:

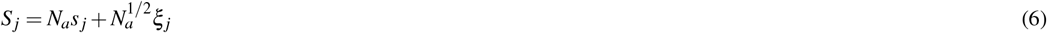

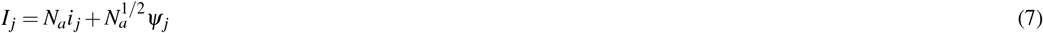

leading to Jacobian and diffusion matrices which can be written as block matrices of the forms:

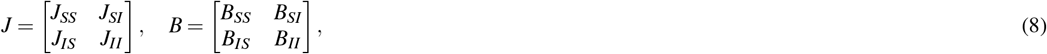

where each submatrix is given in Supplementary Section S2 and the full derivation given in Rozhnova *et al*.^24^. To find the reproductive ratio, *R*_*t*_, over time, we can use the next generation matrix (NGM, *K*),^28,29^, and take *R*_*t*_ = *ρ*(*K*) = *ρ*(*FV* ^−1^) where *ρ* is the spectral radius and the transmission and transition matrices are given by:

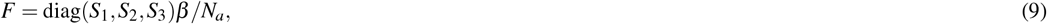

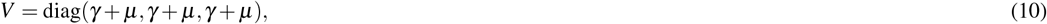

respectively, which results in:

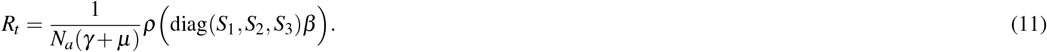

We consider different epidemic transitions caused by changes in the effective contact rate. Such transitions could correspond to the introduction and removal of NPIs and the mutation of a new pathogen variant and aim to represent transitions seen in the COVID-19 dataset analysed in the Applications Section. The baseline model parameters are given in Table 1. These were chosen to give similar dynamics as the Alpha variant of SARS-CoV-2^4,30–32^. Here *β*_0_ is the baseline effective contact matrix.

**Table 1.** Default parameters and values for each set of simulations run. The parameters are chosen to give similar contagion dynamics to the Alpha variant of SARS-CoV-2 in populations of a similar magnitude to Maidstone in the United Kingdom (UK).

| Parameter | Biological Interpretation | Value |
| --- | --- | --- |
| $\beta_0$ | Effective contact matrix rate (baseline) | $\begin{bmatrix} 0.231 & 0.1 & 0 \\ 0.1 & 0.231 & 0.1 \\ 0 & 0.1 & 0.231 \end{bmatrix}$ per day<sup>30–32</sup> |
| $\mu$ | Natural per capita mortality/birth rate | $1/(80 * 365)$ per day |
| $1/\gamma$ | Infectious period | 10 days<sup>33,34</sup> |
| $I_0$ | Initial number infectious in each city | (100,100,100) |

We model epidemic transitions by modifying *β* during the course of our outbreak and changing the initial conditions to simulate local NPIs or new variants as in Table 2, where *H*(*t* − *τ*) is the Heaviside function. More information on the simulation methodology is provided in the Methods Section.

**Table 2.** Changes in the initial conditions and effective transmission matrix for each set of simulations run.

| Effective contact matrix |  |  |  | Initial Conditions |  |
| --- | --- | --- | --- | --- | --- |
| Seeding only in City 3 (b) |  |  |  |  |  |
| $\beta(t) = \beta_0$ | | | | $(I_1(0), I_2(0), I_3(0)) = (0, 0, 100)$ | |
| Reduced spread between cities (c) |  |  |  |  |  |
| $\beta(t) = \beta_0 + H(t - 15)$ | $\begin{bmatrix} 0 & -0.1 & 0 \\ -0.1 & 0 & -0.1 \\ 0 & -0.1 & 0 \end{bmatrix}$ | | | $I_0$ | |
| Reduced spread in City 2 (d) |  |  |  |  |  |
| $\beta(t) = \beta_0 + H(t - 5)$ | $\begin{bmatrix} 0 & 0 & 0 \\ 0 & -0.1 & 0 \\ 0 & 0 & 0 \end{bmatrix}$ | | | $I_0$ | |
| New variant (e) |  |  |  |  |  |
| $\beta(t) =$ | $\begin{bmatrix} 0.05 & 0.025 & 0 \\ 0.025 & 0.05 & 0.025 \\ 0 & 0.025 & 0.05 \end{bmatrix}$ | $+ H(t - 15)$ | $\begin{bmatrix} 0 & 0 & 0 \\ 0 & 0 & 0.00125 \\ 0 & 0 & 0.005 \end{bmatrix}$ | $I_0$ | |
| Reduced spread between cities |  |  |  |  |  |
| $\beta(t) = \beta_0 + H(t - 5)$ | $\begin{bmatrix} 0 & 0 & 0 \\ 0 & -0.1 & 0 \\ 0 & 0 & 0 \end{bmatrix}$ | | | $I_0$ | |

These scenarios are also depicted graphically in Fig. 1

**Figure 1.**
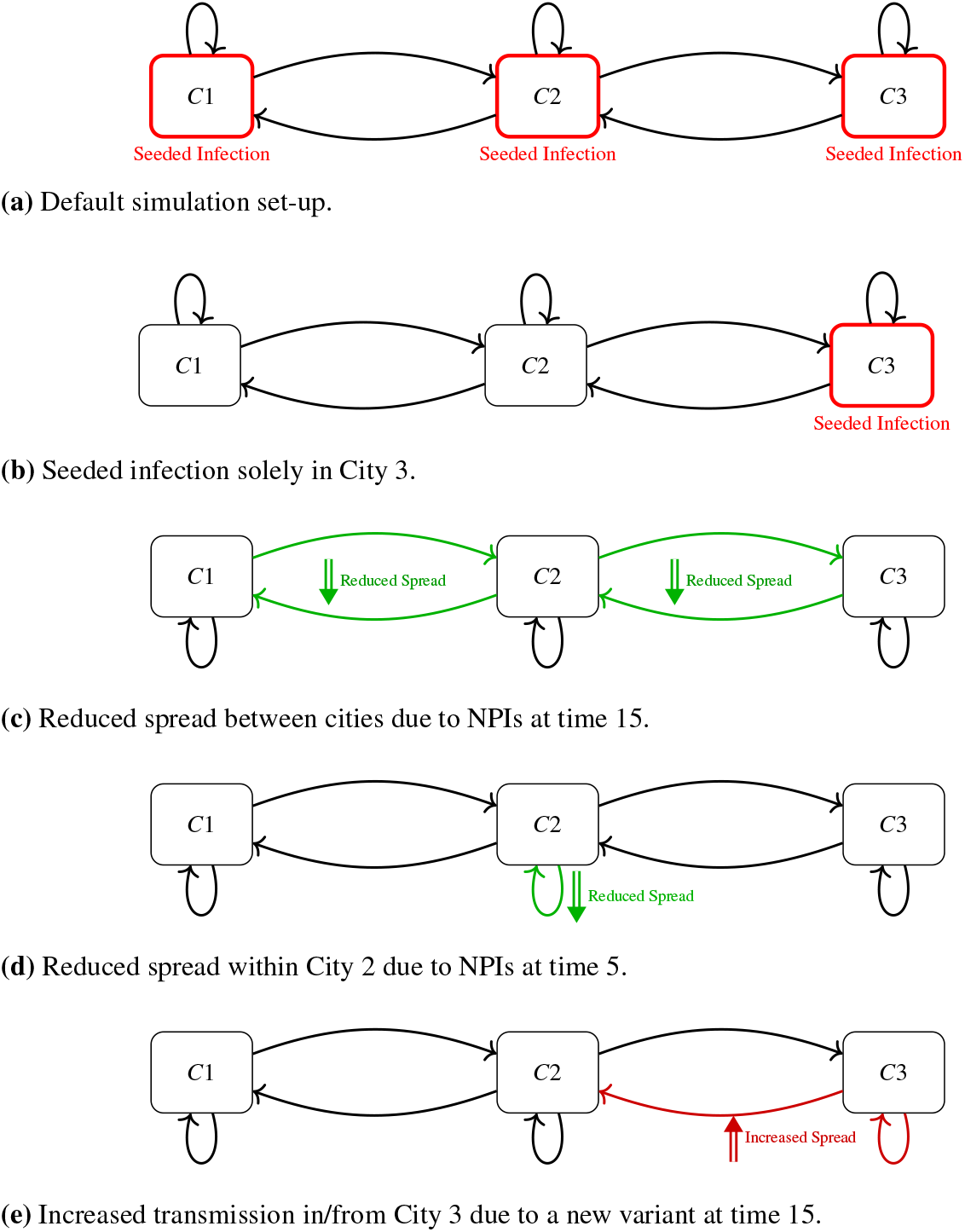
Network diagrams depicting the different modelling scenarios simulated. Each scenario has three cities, each connected to its neighbour.

The theoretical covariance eigendecomposition compares very well to simulations for an epidemic wave with constant effective transmission rate (Fig. 2a). There are two obvious increases in the dominant eigenvalue: on approach to the epidemic peak; and directly afterwards (Fig. 2a, row 3). This mirrors our previous results for a susceptible-exposed-infectious-recovered model with no spatial structure, and confirms that variance-based EWSs can provide evidence, both that the peak of an epidemic is approaching, or has been passed^4^. The normalised eigenvalue also increases on approach to the transition, as expected from the literature^11^. Notably, there is a large spike in this statistic at the *R*_*t*_ = 1 transition at the epidemic peak. This may also provide an extra indication that the outbreak is approaching either elimination or endemicity (i.e. it has gone through its peak). Looking at the dominant eigenvector, it seems to provide information on the spreading patterns of the pathogen. During the early growth phase of the infection waves, the three cities have similar prevalence levels. However, the part of the dominant eigenvector corresponding to City 1 is much larger than that for Cities 2 and 3. Further, Cities 2 and 3 have identical eigenvector elements. This mirrors the infection dynamics, with City 1 having a higher peak and Cities 2 and 3 showing identical dynamics. The dominant eigenvector also shows a dramatic shift at the epidemic transition, possibly corresponding to a rotation of the eigenbasis to account for the differing infection dynamics after the peak. In all sets of results the covariance statistics calculated from the simulations closely match the theoretical results. Small numerical deviations can be seen near the end of the simulations which is expected due to the small number of infectious individuals remaining in the simulations.

**Figure 2.**
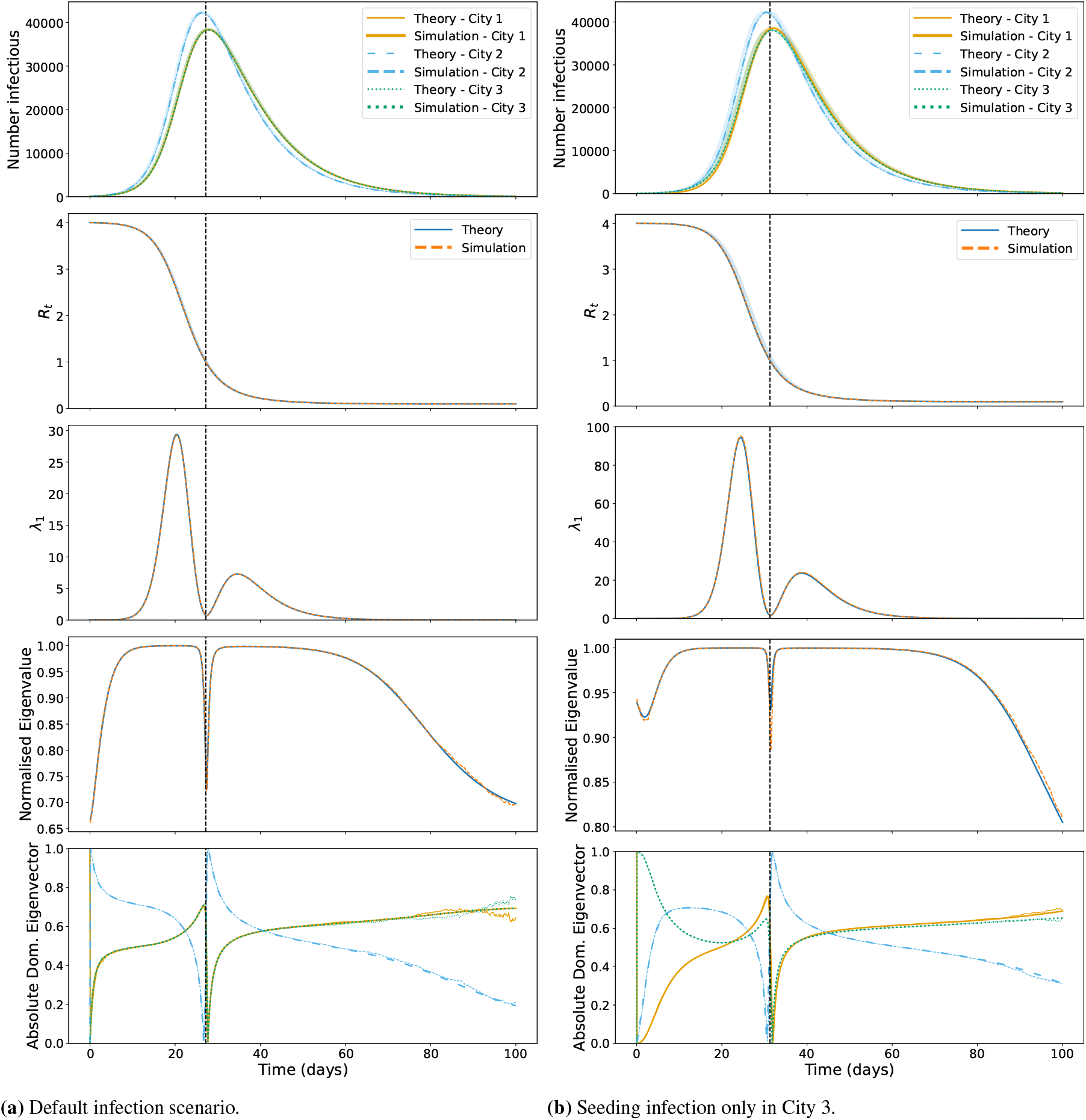
Outbreak dynamics and associated metrics. **Top row** Infectious individuals in each city. **Second row** *R*_*t*_ across the time period. **Third row** Dominant eigenvalue of the covariance matrix of the three cities. **Fourth row** 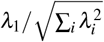. **Bottom row** Absolute dominant eigenvector. Simulation results correspond to 100,000 realisations of the stochastic model with the mean plotted and the 95% prediction interval shown by the shaded area. **Left** Default infection scenario results. **Right** Results from seeding infection in City 3 only. The dashed vertical lines represent the epidemic transitions.

Comparing the above results to those in the scenario where we seeded infections only in City 3 (Fig. 2b), we can immediately see a different trend in the normalised eigenvalue and absolute dominant eigenvector. Although the infection dynamics are similar between the scenarios, the normalised eigenvalue dips before following a similar pattern. This may be due to slower infection take-off compared to Fig. 2a. The dominant eigenvector again provides extra information on the infection dynamics; although infection is lower in City 2 to start with, its element of the dominant eigenvector soon becomes larger than for City 2 and City 3, possibly providing advanced warning of its outbreak size before the difference in prevalence curves is noticeable.

The next set of results corresponds to scenarios modelling the restriction of transmission between cities and within City 2 only (Fig. 3). We again see the distinct two-peak pattern to the dominant eigenvalue, centred around the peak of the epidemic, although this is shifted when transmission is decreased between cities (likely due to the widened peak of infection). However, there is a clear change in gradient of the *λ*_1_ curve when the transmission is decreased between cities, which may help to signal a change in infection rate (though this was visible in the infection curves too).

**Figure 3.**
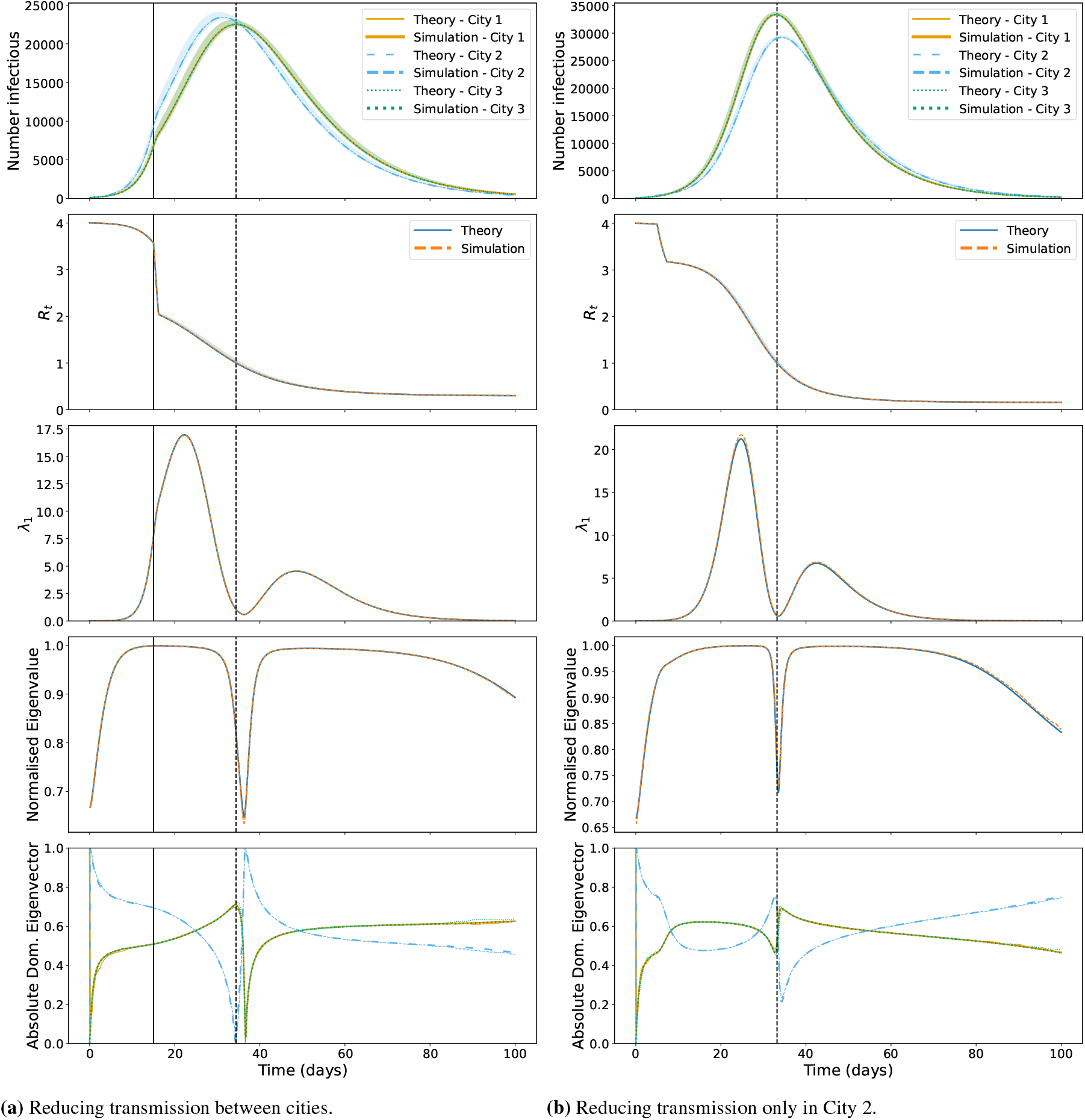
Outbreak dynamics and associated metrics. **Top row** Infectious individuals in each city. **Second row** *R*_*t*_ across the time period. **Third row** Dominant eigenvalue of the covariance matrix of the three cities. **Fourth row** 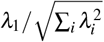. **Bottom row** Absolute dominant eigenvector. Simulation results correspond to 100,000 realisations of the stochastic model with the mean plotted and the 95% prediction interval shown by the shaded area. **Left** Results from reducing transmission between cities. **Right** Results from reducing transmission in City 2 only. The solid vertical lines indicate where transmission reduction begins and the dashed vertical lines represent the epidemic transitions.

In both scenarios, the dominant eigenvector looks the same as in our baseline scenario until the transmission rate is reduced (as prior to this, it follows the same dynamics as the baseline scenario). When transmission is reduced between cities, the dominant eigenvector remains qualitatively similar to the baseline scenario (as transmission is higher within a city than between cities). For the scenario where transmission within City 2 is decreased however, there is a clear decrease in the element corresponding to City 2 pre-empting its now smaller peak compared to Cities 1 and 2. This reinforces that the dominant eigenvector can provide extra information on the infection dynamics of the system.

Finally, we modelled another scenario where the system is tending towards elimination before introducing a more transmissible variant in City 3 (see Fig. 4). As expected, there is similar behaviour in *λ*_1_ and the normalised eigenvalues to the other simulations and the dominant eigenvector behaves the same as the baseline until the point transmission is increased from City 3. It is clear, however, especially when considering Fig. 4b that there is a subtle shift in the gradient of *λ*_1_ and the normalised eigenvalue, and a much more visible rotation in the dominant eigenvector on approach to the re-emergence transition. This is a good sign that our proposed statistics could work as both a signal of an upcoming peak and as an EWS of a critical transition. The increased simulation variance compared to the other modelled scenarios is because delayed ‘take-off’ of the epidemic wave leads to greater variability in the observed epidemic curves.

**Figure 4.**
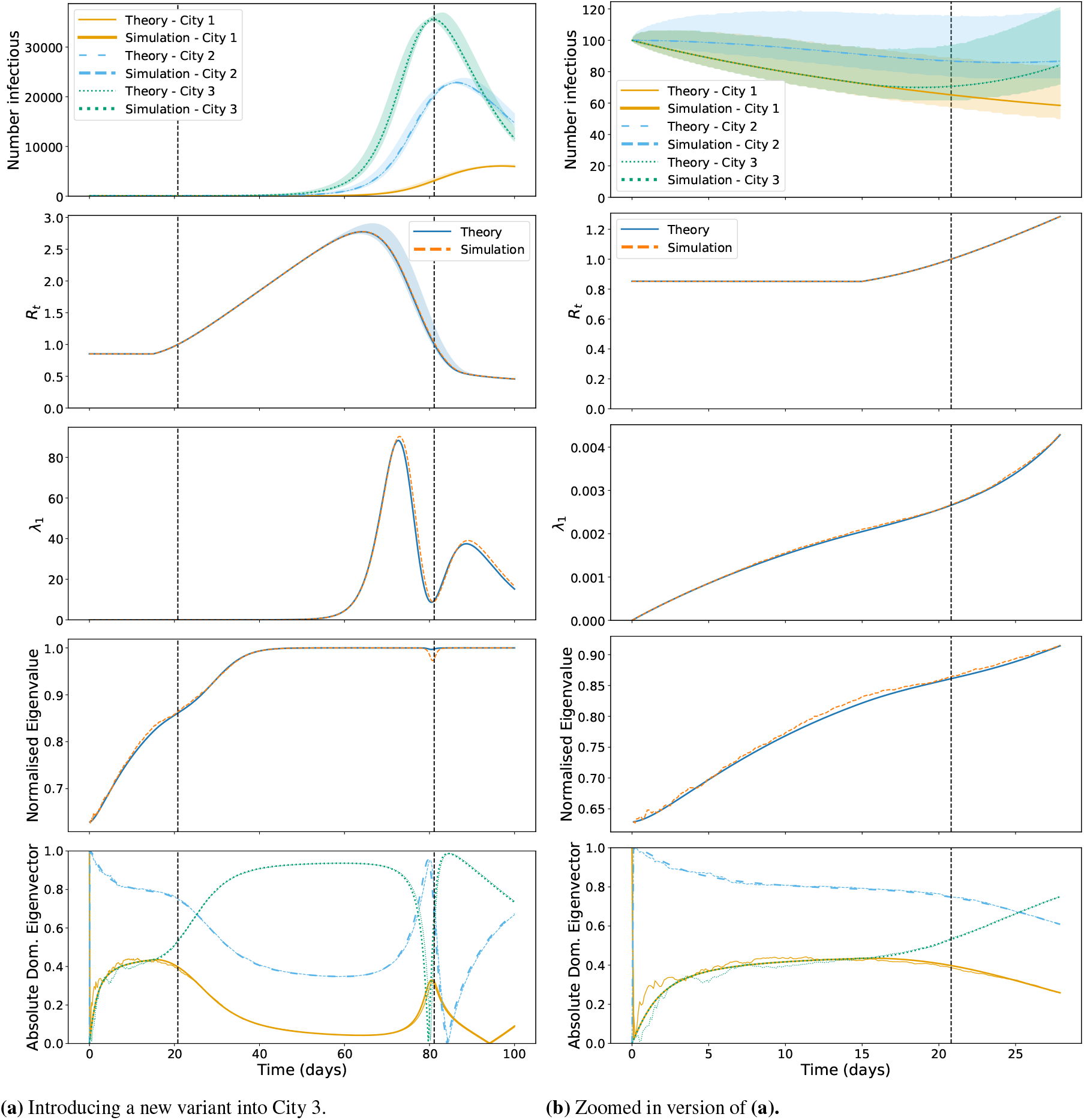
Outbreak dynamics and associated metrics. **Top row** Infectious individuals in each city. **Second row** *R*_*t*_ across the time period. **Third row** Dominant eigenvalue of the covariance matrix of the three cities. **Fourth row** 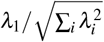. **Bottom row** Absolute dominant eigenvector. Simulation results correspond to 100,000 realisations of the stochastic model with the mean plotted and the 95% prediction interval shown by the shaded area. **Left** Results from introducing a new variant into City 3. **Right** Zoomed-in version of left for days 0 to 30. The solid vertical lines indicate the introduction of the new variant and the dashed vertical lines represent the epidemic transitions.

In addition to the above simulation results, we also reran each scenario with a 10-fold decrease in population size to 10,000 individuals per city to investigate the effect on the eigenbasis trends. With the 100,000 individuals present in each city, we are operating in a macroscopic regime where the stochastic simulations closely match the ODE results. However, with a decreased population size we are likely in a ‘mesoscopic’ system size wherein there is more variance between simulation results. This can indeed be seen in the results in Supplementary Section S1.1. Nonetheless, the theoretical and simulation-based dominant eigenvalues show good concordance across all scenarios simulated. Further, although the simulation-based dominant eigenvectors have more numerical instability than those with 100,000 individuals in each city, they still have high concordance to the theoretical values with the same qualitative trends present.

As real-world infectious disease data generally deals with case counts (incidence) as opposed to prevalence (the number of infectious individuals) and we have previously shown that EWS calculated on incidence data can show qualitatively different trends to those predicted by theoretical prevalence results^4,35^, we also tested whether using incidence or cumulative cases impacts the observed eigenbasis trends (see Supplementary Section S1.2). Interestingly, the eigenbasis of the incidence covariance matrix behaves identically to the prevalence. The eigenbasis of the cumulative cases, however, is less reactive to changes in *R*_*t*_. Although the eigenvectors in the beginning of the outbreak show a similar relationship to the final size of the epidemic as with prevalence and incidence, the dominant eigenvalue has only one temporal mode centred on the *R*_*t*_ = 1 epidemic transition (which may suggest the dominant eigenvalue could be used as confirmation that the epidemic peak has passed). Further, the normalised eigenvalue is flat throughout most of the epidemic wave and so does not hold much predictive power. Nevertheless, these results suggest that covariance matrices calculated on cases also capture the underlying Jacobian dynamics of an infectious disease system.

### Application: SARS-Cov-2 in England

The SARS-CoV-2 pandemic has provided a previously unprecedented quantity and quality of infectious disease case data. With high granularity in both age and location of infected individuals, it provides an ideal dataset to test the proposed EWSs. As previously analysed in the literature, EWSs successfully anticipated the peaks and troughs in the case data of various countries^4,6,7,36–38^. However, multivariate analysis such as our proposed EWSs have not, to our knowledge, been tested on case data. We therefore consider whether the eigendecomposition of both the spatially-structured and age-structured covariance matrices provide EWSs of the epidemic transitions in the United Kingdom Health Security Authority (UKHSA) case data. This dataset was chosen as it provides a high level of spatial granularity in case data (the data was previously analysed in Looker *et al*.^4^).

To begin, we undertake a similar pre-processing and detrending method as in Looker *et al*.^4^, before considering the eigenvalues and eigenvectors of the rolling covariance matrix in age- and spatially-structured case incidence and their corresponding detrended time series. See the Methods Section for more information on the methods used.

#### Age-stratified data

We first look at the age-stratified case data of Maidstone, an area in the United Kingdom. It is a lower tier local authority (LTLA), the smallest-sized area for which the UKHSA published case data and so can be used to explore the effects of spatial aggregation on our results. We have already successfully analysed Maidstone using variance-based EWSs^4^. We consider the 2020–2021 period of the pandemic due to the number of epidemic transitions present in the data.

Figure 5 displays the incidence and rolling mean of the COVID-19 cases in Maidstone with the epidemic transitions overlaid as vertical lines. The corresponding EWSs are calculated based on the age-structured Maidstone case data aggregated into 10-year age classes (0–10, 10–20, … ). We can observe near-monotonic increases in the magnitude of the dominant eigenvalue on approach to the epidemic transitions, especially those occurring in the Christmas 2020 period. Estimating the covariance on a larger rolling window size smooths out the dominant eigenvalue, though the same qualitative trends are present. For the normalised eigenvalue, there are clear spikes on approach to the peaks. This behaviour differs from the theory and simulations (where the spike occurred at the epidemic peak), but likely corresponds to where *R*_*t*_ has increased above one and could be because the approach to the peak introduces new dynamics changing the covariance eigenmodes. There are similar trends between the statistics calculated on the incidence and detrended time series, with the notable difference being the sharpness and frequency of the spikes in the detrended results. This may be because the raw case data has other sources of stochasticity in addition to infection dynamics that are clouding the signals.

**Figure 5.**
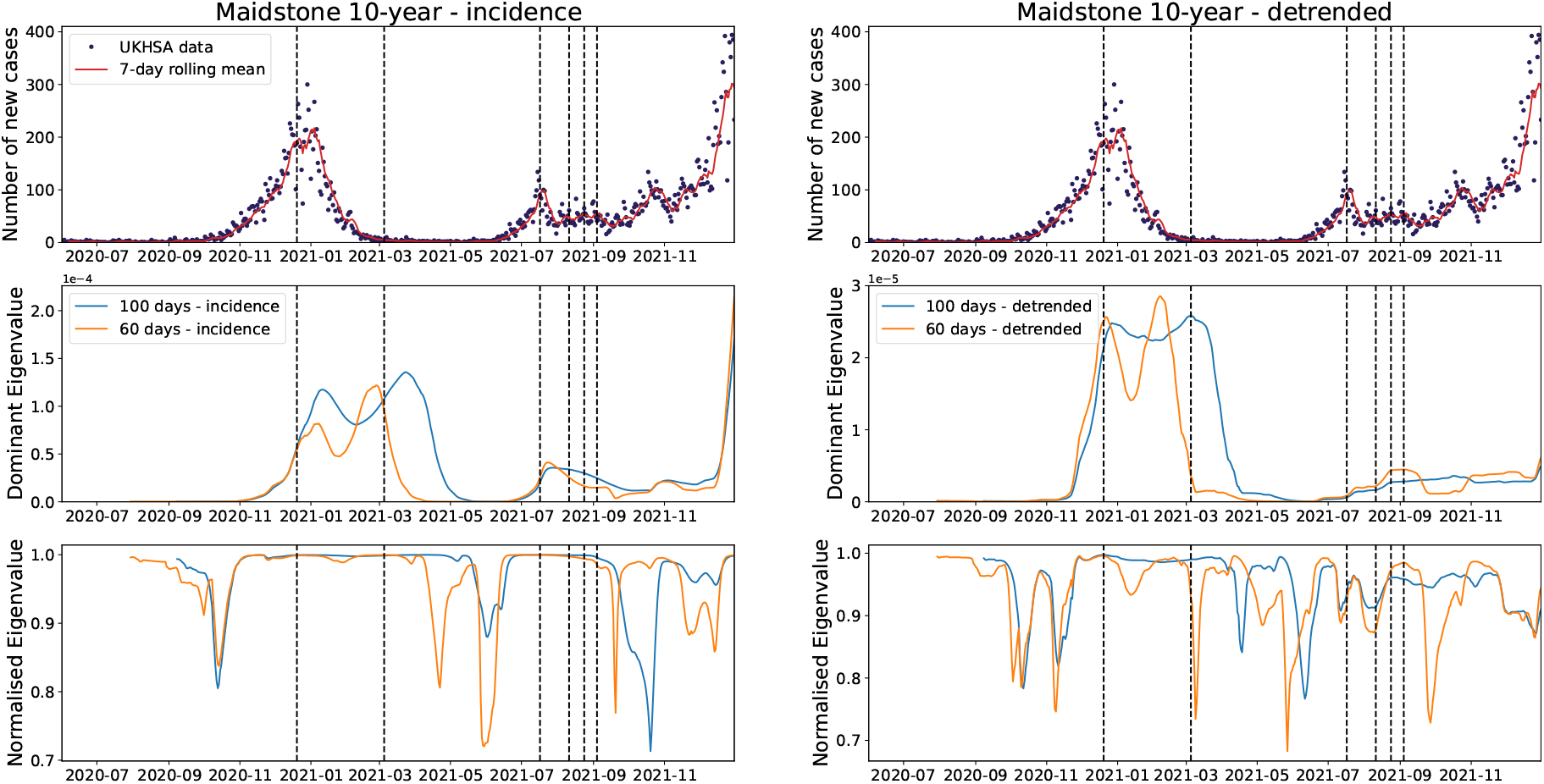
**Top** Incidence and rolling mean of COVID-19 cases in Maidstone. **Middle** Dominant eigenvalue of the covariance matrix of the 10-year age-aggregated incidence (**left**) and detrended (**right**) time series using a rolling window of 100 and 60 days. **Bottom** 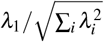.

It is worth noting that the qualitative behaviour of these statistics calculated on the 10-year age-class data is near-identical to the corresponding statistics conducted during a sensitivity analysis using 5-year age-class data in Supplementary Section S1.3. The main difference being that the dominant eigenvalue is nearly half the magnitude of that using the 5-year age class likely due to the decreased stochasticity from further aggregating the data, which is analysed in Supplementary Section S3.

We continue by looking at the behaviour of the dominant eigenvector of the empirical covariance matrix for the same dataset. As seen in Fig. 6, there are periods of large oscillations before many of the epidemic transitions, especially those corresponding to (re-)emergence. Conversely, when the cases time series is going through an infection wave (such as the Christmas 2020 peak) the eigenvectors are near-stationary and this is likely because the stochasticity is dominated by the infection dynamics and has reached a quasi-stationary distribution. The results are then similar to those expected from the literature, wherein the eigenvector is constant^11,19^. Between the results calculated on the incidence and detrended time series, most of the qualitative behaviour is the same. However, the elements corresponding to the 90+ and 10–19 age-classes have different trends on approach to the re-emergence in July 2021. This is likely due to the small number of infections. Nevertheless, rapid changes in the dominant eigenvector are present in both the incidence and detrended results on approach to the July 2021 peak and so may have provided early warning of the upcoming infection wave. Similar trends are seen when increasing the rolling window size from 60 to 100 days as with the eigenvalues, namely that there is less noise in the trends displayed by the eigenvectors and that they stay near-constant for longer.

**Figure 6.**
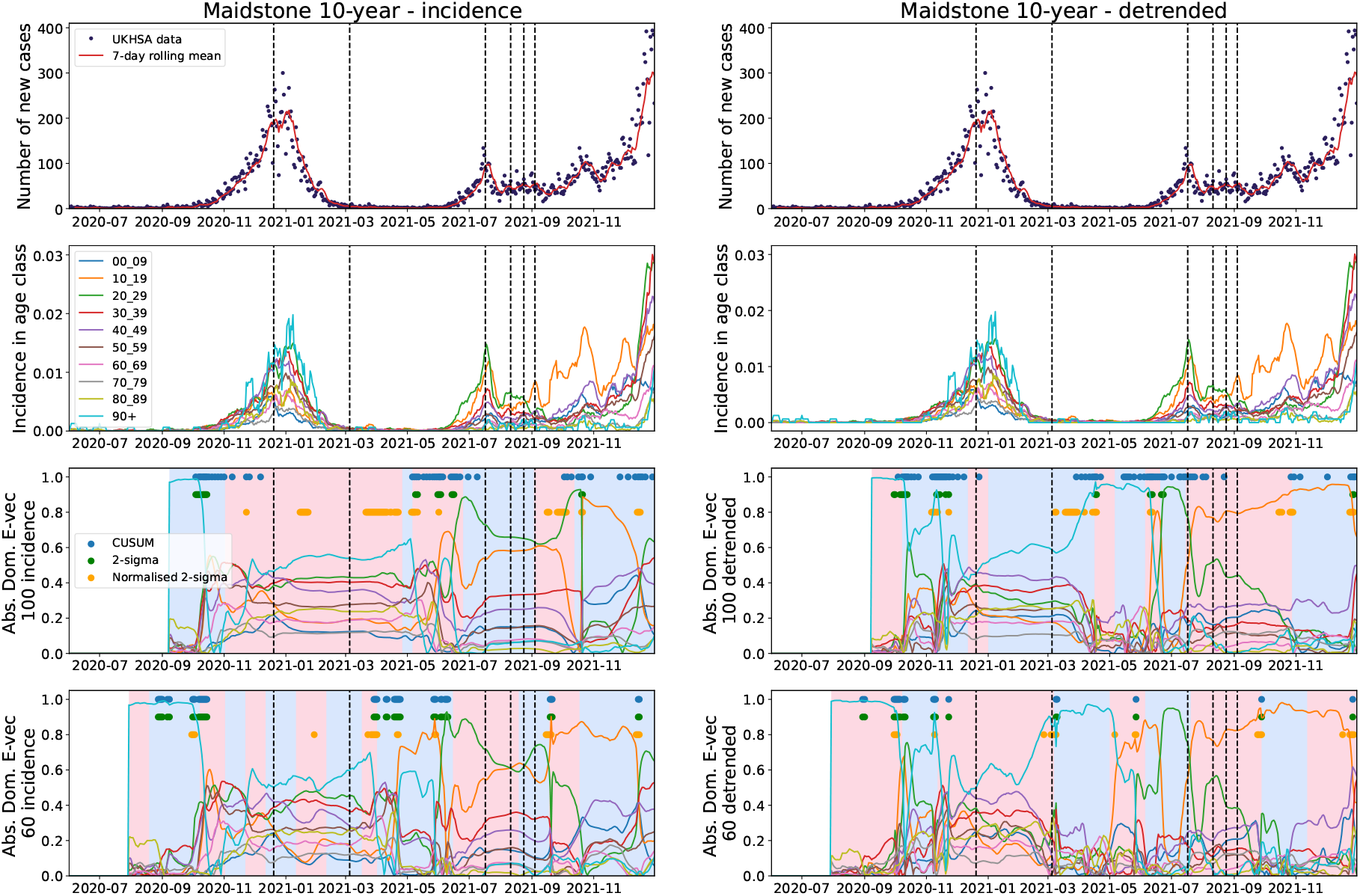
**Top row** Incidence and rolling mean of COVID-19 cases in Maidstone. **Second row** Incidence time series for each age-class. **Third row** Absolute value of the dominant eigenvector of the covariance matrix of the 10-year age-aggregated incidence (**left**) and detrended (**right**) time series using a rolling window of 100 days. **Bottom** Absolute value of the dominant eigenvector of the covariance matrix of the 10-year age-aggregated incidence (**left**) and detrended (**right**) time series using a rolling window of 60 days. Dashed vertical lines show epidemic transitions, solid dots indicate where each method signals a change in the angular rotation and background colour changes are change-points identified by linearly penalised segmentation.

The change in the elements of the dominant eigenvector for younger age classes around June 2021 is likely caused by the re-opening of schools and of enforced lateral flow testing for school children. This reinforces the simulation-based results, where the dominant eigenvector provides information on the nature of the underlying infection dynamics and case statistics. Keeping age-structured data may help analysts to understand trends in case data and provide extra information through EWSs.

Finally, we consider whether qualitative shifts in the dominant eigenvector could be quantitatively detected by a range of statistical tests. Fig. 6 shows a range of online (real-time) change-point detection methods (CUSUM, 2-sigma and normalised 2-sigma) applied to the dominant eigenvector of the covariance matrix, calculated on the incidence and detrended time series, as well as the change points detected by an offline likelihood method (linearly penalised segmentation, shown by the changes in background colour). For more information on each of the tests used, see the Methods Section. In both the incidence and detrended results, we can see that all methods reliably signal changes in the eigenvectors, though the CUSUM and normalised 2-*σ* methods signal more than the 2-*σ* method applied to the raw angle displacement time series. While we think it is still important to investigate the eigenvector behaviour qualitatively, especially due to the extra information it may provide about infection dynamics, this does suggest that the eigenvector can be used as a quantitative EWS too.

We conducted another sensitivity analysis in Supplementary Section S1.4 considering the effects of using a 30-day rolling window when calculating the empirical covariance matrix. The dominant eigenvalue displayed similar behaviour to the 60-day rolling window, but with much more stochasticity and roughness in the normalised eigenvalue. The eigenvectors displayed far more random, oscillatory behaviour with none of the stationary behaviour around the epidemic peaks seen in the 60-day and 100-day results.

#### Age-stratified data: Higher aggregation

As previously stated, it is common to aggregate disease data from smaller sub-populations to larger spatial scales when modelling infectious disease dynamics. In this section, we investigate the effects of spatial aggregation on our proposed EWSs by applying the methods to the National Health Service (NHS) region corresponding to Maidstone, the South East of England (comprising 67 LTLAs, including Maidstone).

Fig. 7 shows that the dominant eigenvalue has much smaller peaks when using the covariance matrix calculated on the aggregated incidence data. There is barely an increase on approach to the first, smaller peak in November 2020 before we again see the similar double spike in the dominant eigenvalue around the January 2021 peak as in the Maidstone results. The dominant eigenvalue corresponding to the covariance matrix of the detrended data however, shows a clear, anticipatory increase on approach to the smaller November peak and so may have provided an EWS. This suggests that for higher aggregation levels it is important to detrend the time series when calculating early warning statistics to regain some of the stochasticity lost from aggregation. This loss of information to aggregation is analysed in Supplementary Section S3. Unlike the Maidstone results, there are large differences in the behaviour of the normalised eigenvalue when using a 100 day rolling window compared to a 60, with almost all the spikes precluding a epidemic transition smoothed out.

**Figure 7.**
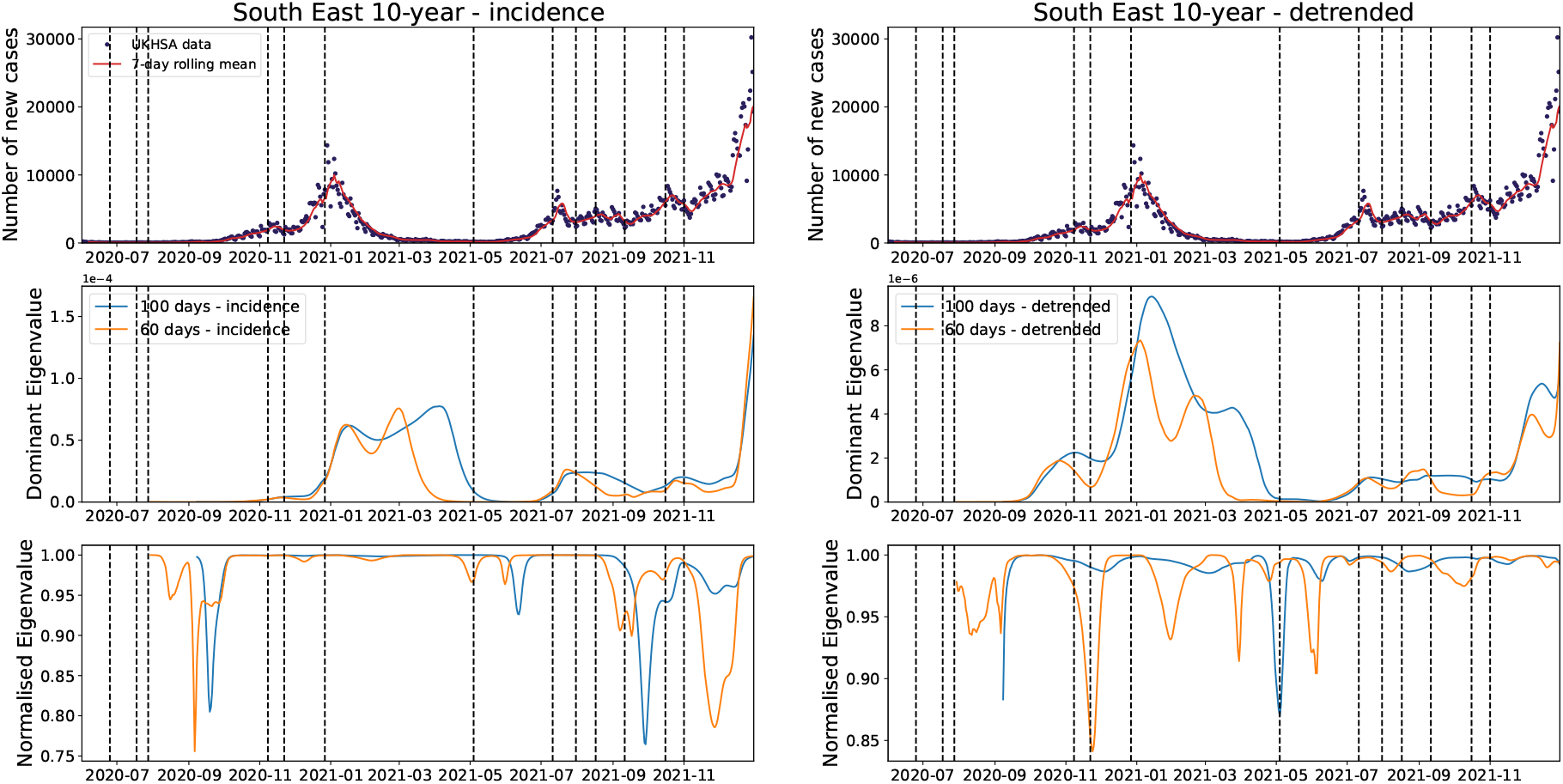
**Top** Incidence and rolling mean of COVID-19 cases in the South East of England. **Middle** Dominant eigenvalue of the covariance matrix of the 10-year age-aggregated incidence (**left**) and detrended (**right**) time series using a rolling window of 100 and 60 days. **Bottom** 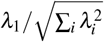.

Similarly to Maidstone, oscillatory behaviour interspersing the long-term stability is also seen on approach to epidemic transitions in the dominant eigenvector in Fig. 8. The largest components of the South East eigenvector however, do differ from that in Maidstone LTLA which may be due to the larger South East area having a larger proportion of working-age individuals than Maidstone, such that they contribute more to the infection dynamics. Finally, the change-point detection results for the South East are similar to those in Maidstone LTLA, although the normalised 2-*σ* method now seems to perform better on the incidence data, with all methods performing well on the detrended data.

**Figure 8.**
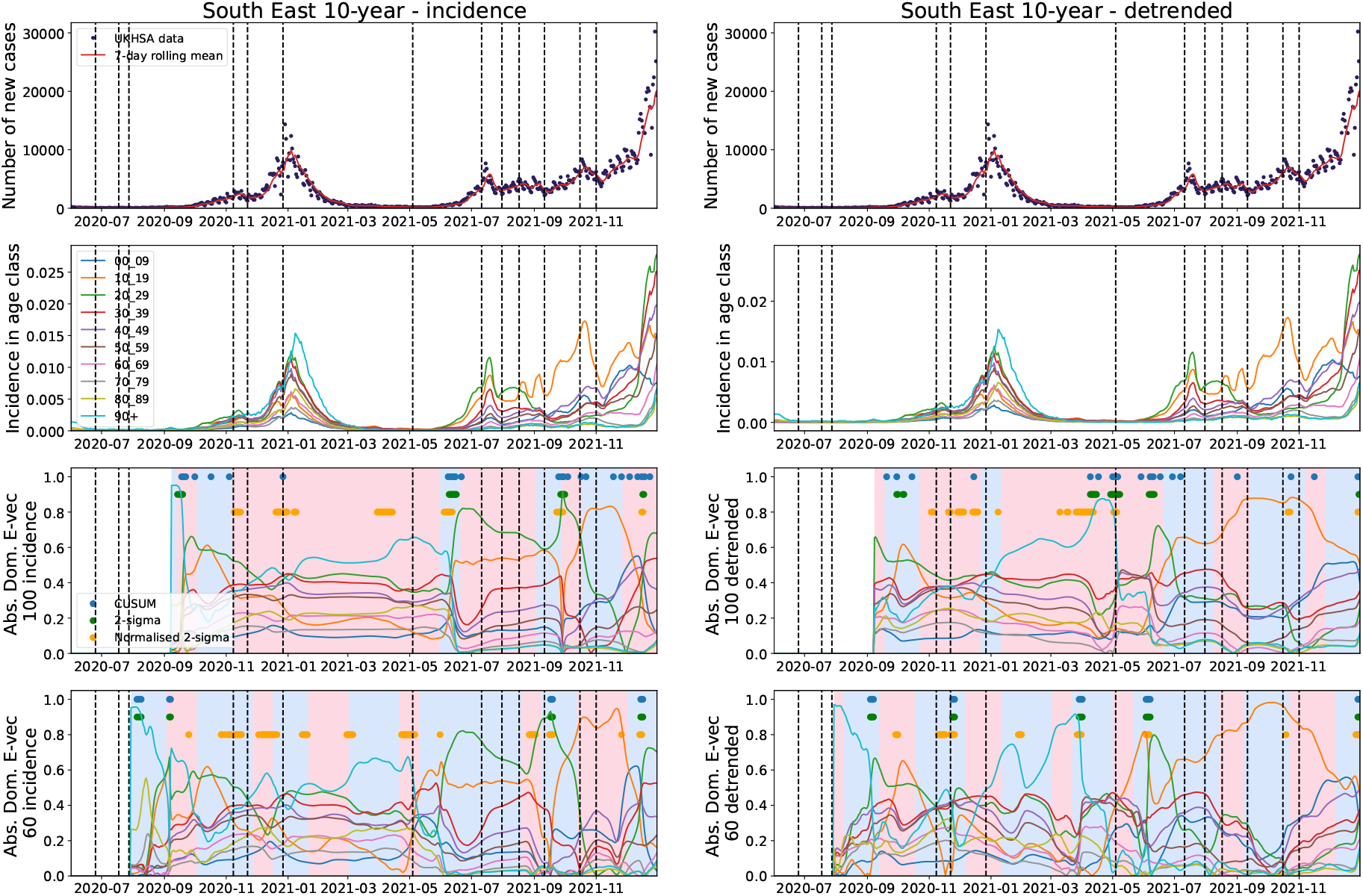
**Top row** Incidence and rolling mean of COVID-19 cases in the South East of England. **Second row** Incidence time series for each age-class. **Third row** Absolute value of the dominant eigenvector of the covariance matrix of the 10-year age-aggregated incidence (**left**) and detrended (**right**) time series using a rolling window of 100 days. **Bottom** Absolute value of the dominant eigenvector of the covariance matrix of the 10-year age-aggregated incidence (**left**) and detrended (**right**) time series using a rolling window of 60 days. Dashed vertical lines show epidemic transitions, solid dots indicate where each method signals a change in the angular rotation and background colour changes are change-points identified by linearly penalised segmentation.

#### Spatially-stratified data

Having observed the predictive power of our proposed EWSs on age-structured disease data, we now consider the applicability of the methods for anticipating epidemic transitions in spatially-structured data. Analysing the total case data across all NHS regions in England, we investigate whether the eigenvalues and dominant eigenvector of the spatial covariance matrix can anticipate the epidemic transitions in the England cases time series.

Figure 9 again shows similar patterns in the dominant eigenvalue on approach to the Christmas 2020 and July 2021 infection waves. However, there seems to be a delayed signal when compared to the timing of the eigenvalue peaks in the Maidstone and South East results. This delayed increase is possibly caused by the spatial spread of epidemic waves through the country^4^. Due to the high level of aggregation, there may also be less of a signal from the smoothing of stochasticity. This may also be why the dominant eigenvalue from the detrended time series peaks slightly earlier and higher (as it is more susceptible to the stochastic fluctuations).

**Figure 9.**
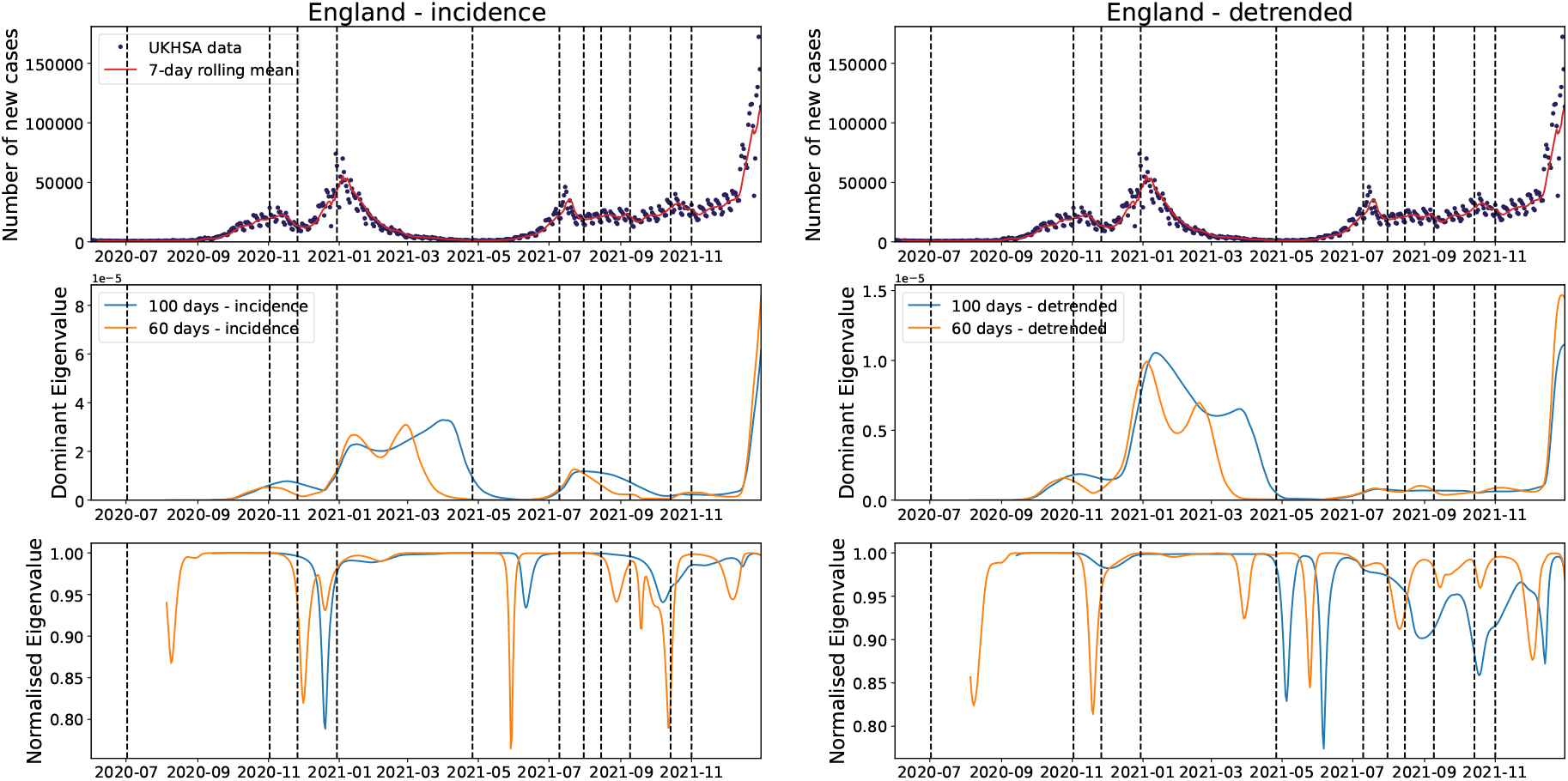
**Top** Incidence and rolling mean of COVID-19 cases in England. **Middle** Dominant eigenvalue of the covariance matrix of the NHS regional incidence (**left**) and detrended (**right**) time series using a rolling window of 100 and 60 days. **Bottom** 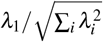.

There is a clear spatial pattern present in the dominant eigenvector shown in Fig. 10 for the NHS regional covariance. The North West region and London have the highest magnitude component of the dominant eigenvector for a large proportion of the pandemic period analysed. This may be due to the North West becoming the epicentre of the Delta variant in England^39–41^. Around the Christmas 2020 peak, London, the East and South East regions shift to becoming the main components of the eigenvector in the 60-day rolling window results. This could be due to the Alpha variant which likely originated near here, which has also been signalled by other variance-based EWS^4^. This is shown to a lesser extent in the 100-day results, where the North East and Yorkshire also have a large component. Nonetheless, these results again suggest that the covariance eigenvectors may provide further insight into the spreading dynamics of the pathogen. Finally, the online detection methods perform similarly to the Maidstone LTLA and South East results.

**Figure 10.**
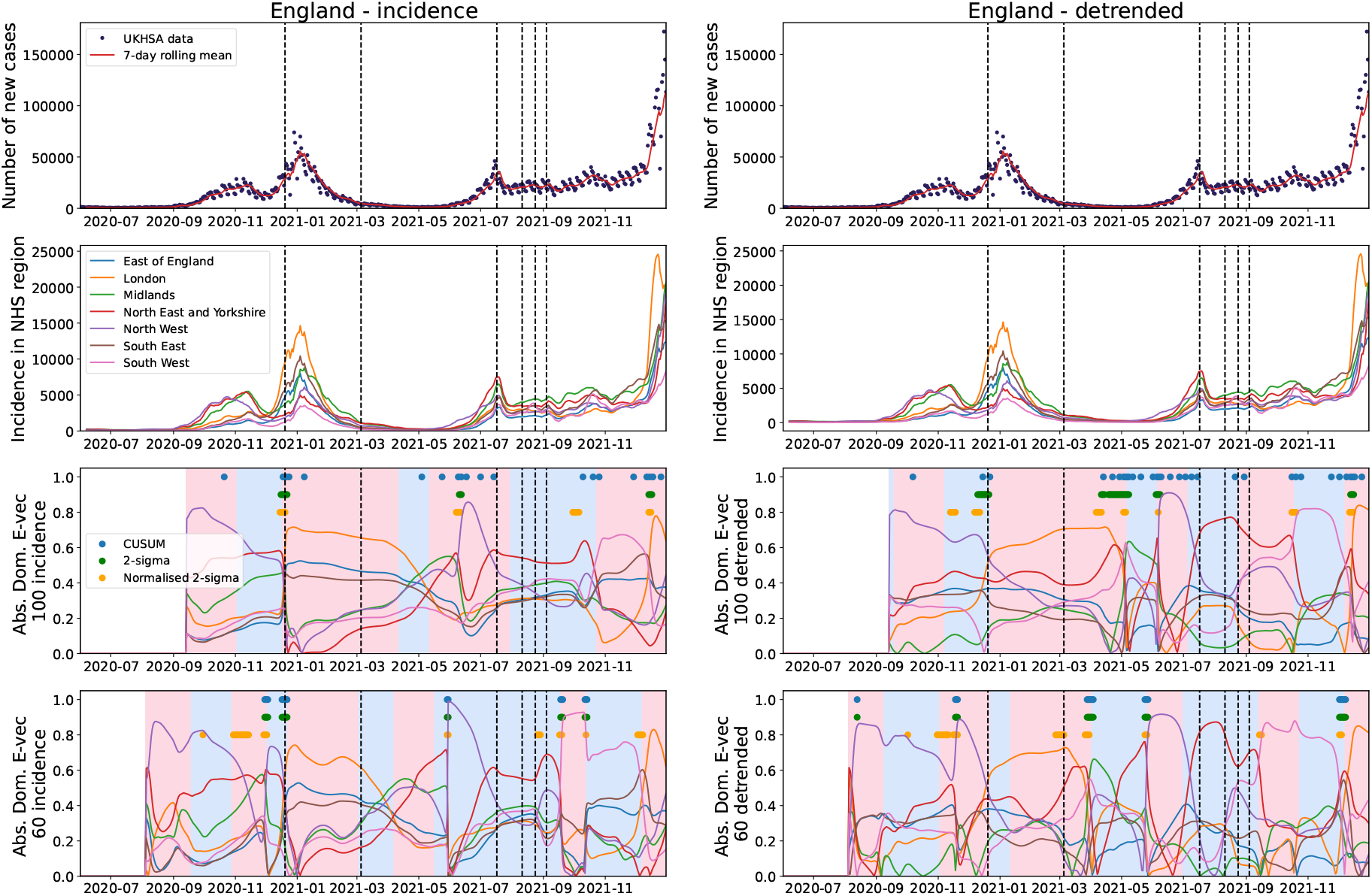
**Top row** Incidence and rolling mean of COVID-19 cases in England. **Second row** Incidence time series for each NHS region. **Third row** Absolute value of the dominant eigenvector of the covariance matrix of the NHS regional incidence (**left**) and detrended (**right**) time series using a rolling window of 100 days. **Bottom** Absolute value of the dominant eigenvector of the covariance matrix of the 10-year age-aggregated incidence (**left**) and detrended (**right**) time series using a rolling window of 60 days. Dashed vertical lines show epidemic transitions, solid dots indicate where each method signals a change in the angular rotation and background colour changes are change-points identified by linearly penalised segmentation.

## Discussion

This paper analyses the theoretical evolution of the covariance matrix of linearised metapopulation and age-structured infection dynamics and the ability of related EWSs to anticipate epidemic transitions. We investigated the trends in the dominant eigenvalue and eigenvector through a mathematical and simulation-based study. Our analysis considered both age- and spatially-structured reported COVID-19 case data in England from June 2020 to December 2021. In addition, we analysed the impacts of data aggregation on observed early warning statistics. Finally, we also investigated whether online change-point detection methods could detect changes in the covariance matrix eigenbasis.

We built on existing results in the literature indicating the usefulness of covariance-based statistics in an EWS context^10,13,15–18^. By focusing on the dominant eigenvector of the covariance matrix, we removed some of the requirements of previous studies, such as requiring knowledge of community structure in ecological models^16^ or both the growth rate and abundance in cell biology^13^, to find a generalised relationship with the Jacobian dynamics. The simulation results show that the dominant eigenvector can provide infectious disease modellers with additional information on the spreading patterns of pathogens even with low incidence or at the very beginning of an outbreak. These findings are reinforced by the COVID-19 results which indicate that EWS calculated on the covariance matrix might have provided advanced warning of the second peak in December 2020 and the arrival of the Alpha and Delta variants into the UK. Future work should analyse the covariance matrix of other case datasets to see if similar results around the infection dynamics can be identified.

The successful online detection of the change-points in the covariance eigenbasis indicates its applicability to signal changes in the infection dynamics. This ability can also be seen in the dominant eigenvalue and normalised eigenvalue results. However, we note that the sudden shifts in the eigenbasis in the theoretical results are centred on the *R*_*t*_ = 1 transition, which suggests that these methods may also be suited as a confirmation tool of these transitions (such as confirming that an epidemic peak has occurred as opposed to a stochastic drop in cases). Much of the existing literature on theoretical EWS for epidemiology models the statistics of prevalence, with modelling choices such as the reporting distribution required to then extend this to observed incidence^4,35,42^. In our results, we recorded the *I* to *R* transition as cases to account for reporting delays and because individuals are unlikely to test or display symptoms as soon as they become infected. If a different incidence calculation method was used then the covariance eigenbasis of the incidence and prevalence may not have matched, and the peaks in changes in the eigenbasis evolution could anticipate/precede the epidemic transition instead. Modellers should thus consider what impact this will have on observed EWS when using incidence in practice compared to the prevalence theory.

Our comparison between statistics calculated using aggregated and disaggregated case data highlights the benefits to modellers in using disaggregated data where available. Although it adds model and numerical complexity, using more model subclasses provides more information on the infection dynamics. We have also shown that in an EWS context, this extra information can provide insight into spreading dynamics or upcoming transitions that using aggregated subclasses (such as broader age classes or larger spatial areas) does not. Our mathematical analysis in Supplementary Section S3 indicates that these findings can be attributed to the projection of the higher-dimensional dynamics (the disaggregated data) onto a lower-dimensional subspace (the aggregated data). Future work should investigate the trade-off between the greater stochasticity in disaggregated models and the impacts of movement between model subclasses, such as movement between network nodes or age classes, on the observed signals.

It should be noted that estimates of both the eigenbasis and the covariance matrix itself are numerically unstable^11^. With the high frequency and granularity of the data available to us in both our simulation-based and COVID-19 case data-driven studies, this wasn’t problematic. However, for other epidemics applications, there will likely be less data available which may impact the results. In addition, our sensitivity analysis on the rolling window size indicates that observed trends can depend on the window size used. Choosing a too large window size smooths out changes in the covariance eigenbasis that happen on a shorter time scale whilst a too small window size increases noise. The methods also make an inherent assumption of near-ergodicity when calculating the rolling window statistics of the case data. In standard EWS analysis, the covariance matrix and other EWS statistics are calculated near a stationary point such that this assumption is generally satisfied. It is less clear how the non-stationarity of the system impacts the numerical statistics in the non-stationary case. Nonetheless, the applicability of the methods to both the simulations and COVID-19 dataset suggest that both the covariance matrix and the Jacobian vary slowly enough for observed statistics to be representative of the system dynamics.

In this article, we have used both simulation-based and data-driven analyse to investigate the applicability of the eigenbasis of the non-stationary covariance matrix of metapopulation infection models as an EWS. We show that using multivariate statistics in infectious disease modelling can provide extra information and EWS to modellers.

## Methods

All data analysis and simulations were conducted using Python 3.11 with packages: NumPy (version 1.26.4), Matplotlib (version 3.84), SciPy (version 1.13.0), Ruptures (version 1.10.0), epyestim (version 0.1) and Pandas (version 2.2.1). A repository containing the data and code used to conduct this study can be found at https://github.com/joshlooks/covariance-matrix.

### Simulations

Theoretical simulations were conducted using the Gillespie method^43^ with the theoretical covariance eigenbasis evolution calculated using numerical integration. We ran 10,000 simulations for each of the modelling scenarios for city populations of 100,000 and 10,000 individuals. The simulations were ran for a maximum of 100 days. Events and transition rates for the siulations are given in Table 3. In order to account for symptom onset and reporting delays, we took the incidence of cases to be the number of *I* → *R* transitions with an aggregation period of one day to match the COVID-19 reporting period. This could also be considered as an individual being tested upon arrival to hospital. The simulation results were detrended by subtracting the empirical mean of all realisations, and the empirical covariance matrix of these detrended residuals was calculated using *NumPy* which uses an unbiased estimate given as

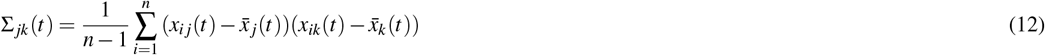

where *n* is the number of simulations, *x*_*i j*_ is the prevalence or incidence in City *j* for simulation *i* and 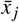 is the average prevalence or incidence in City *j* ith measurement collected at the jth cell. Both the empirical and theoretical eigenbases were then calculated using *NumPy*.

**Table 3.**
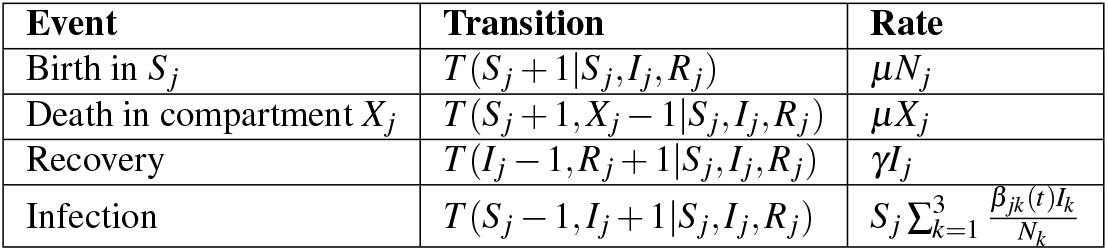
Events and transition rates for the Gillespie simulations. Here *T* (*Y | X* ) indicates a transition from a system state of *X* to a system state of *Y* .

### Early warning signal calculation: Data

Although briefly mentioned in the Applications Section, we provide here a more thorough explanation of the UKHSA COVID-19 dataset and how it was used to estimate the age-structured and spatial-structured covariance matrices. For a full explanation of the dataset and detrending rationale, see the Methods section in Looker, 2025^4^.

For age-structured data, we use the daily reported cases time series from the, now decommissioned, COVID-19 dashboard^44^. This is a combination of lateral flow tests and polymerase chain reaction-only (PCR) tests and specifies the number of cases at the LTLA level, grouped into 5-year age classes. We choose to focus on the Maidstone LTLA and its corresponding NHS region of the South East of England, as previous analyses successfully used other variance-based EWS to anticipate the epidemic transitions in their case data^4^. The Maidstone case data is aggregated up to seven-day rolling sums. We then assume that infection dynamics are similar for all LTLAs in the South East NHS region and detrend the time series within each LTLA by subtracting the corresponding NHS regional average. We also consider the impacts of aggregation on the EWS by taking the resulting age-structured case data for the South East and detrending using the average across all NHS regions in England.

For spatially-structured data, we use PCR case data to reduce the effects of the evolving testing requirements and test availability on the results. The case data is aggregated up to seven-day rolling sums for each NHS region. We detrend these aggregated case counts by subtracting the average over all NHS regions in England.

To calculate the EWSs, we calculate an unbiased empirical, rolling covariance matrix across the age- or spatially-structured case data with a window size of either 60 or 100 days. This was done using the *Pandas* Python library. The eigendecomposition of this matrix was then calculated using the *Numpy* Python library, with a linear-sum assignment used to account for any sign changes in the eigenvectors. Early warning signals were then calculated using this method on both the (normalised) incidence and detrended case data. Each entry of the covariance matrix, Σ, is estimated as

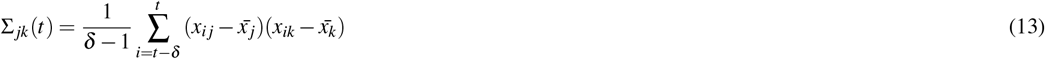

where *δ* is the window size used, *x*_*i j*_ is the incidence in age-class or spatial area *j* for time-point *i* and 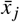 is the average incidence in age-class or spatial area *j*.

### Change-point detection methods

Here we explain the different statistical tests used when quantitatively evaluating the rate of rotation of the dominant eigenvector as a possible EWS of changing infection dynamics. The rate of rotation was quantified using the angular displacement given by

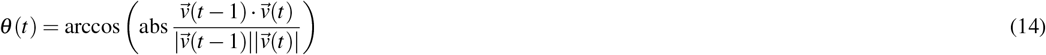

where *θ* (*t*) is the angular displacement, 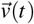 is the dominant eigenvector at time *t* and 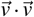 and | · | denote the dot product and vector magnitude, respectively. The absolute value of the cosine similarity was used such that *θ* (*t*) ∈ [0, *π*/2] to compare better with other positive EWS such as the variance.

To provide a baseline for assessing the applicability of changes in the dominant eigenvector as an EWS, we used linearly penalised segmentation^45^ in the *Ruptures* Python library^46^ as an offline change-point detection method. The minimum size and penalty hyperparameters were chosen as (10, 20) for Maidstone 10-year data; (20, 20) for the South East and England level data, apart from the detrended data using a rolling window size of 60 for which we used (20, 30). These were chosen arbitrarily but the identified change-points correspond well to the qualitative changes in the dominant eigenvector.

For the proposed EWS, we tried the 2-*σ* method^47^ due to its previous success in anticipating epidemic transitions on traditional early warning statistics. This was applied to both the raw angular displacement data and the ‘normalised’ angular displacement (calculated by subtracting the rolling mean and dividing by the rolling standard deviation). The other method used is the cusum method^48^ which has been used very successfully in other change-point applications^49^. Change-points are identified using the cumulative sum formula

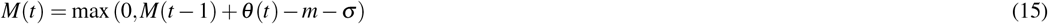

where *M*(*t*) is the cumulative sum of *θ* at time *t, m* = 0 is the assumed mean/drift in *θ* and *σ* is the noise level in the data. The change-points are then identified when *M*(*t*) exceeds a chosen cutoff threshold at which point *M*(*t*) is reset to 0. For the Maidstone 10-year data we set the noise level as (0.01, 0.03) for the rolling window sizes of 100 days and 60 days and thresholds as (0.1, 0.2), as we estimated there would be almost double the amount of noise in the 60 day rolling estimates than in the 100 day. For both the South East and England level data we used (0.2, 0.4) and (0.005, 0.01) for threshold and noise level for the rolling window sizes of 100 days and 60 days respectively.

## Data Availability

A repository containing the data and code used to conduct this study can be found at https://github.com/joshlooks/covariance-matrix. An archived version of this repository is available at https://doi.org/10.5281/zenodo.19678071.

## Acknowledgements

JL is supported by the Engineering and Physical Sciences Research Council through the Mathematics of Systems II Centre for Doctoral Training at the University of Warwick (reference EP/S022244/1). The funders played no role in the study design, data collection and analysis, decision to publish, or preparation of the manuscript.

## Author contributions statement

J.L. conceived the study and wrote the paper. J.L. analysed the data and conducted the computational experiments. All authors analysed the results and reviewed the paper.

