## Supplementary Information for "The covariance matrix of metapopulation disease models and applications to early warning signals"

### Supplementary Materials for The covariance matrix of metapopulation disease models and applications to early warning signals

#### S1 Additional Figures

##### S1.1 Decreased population simulations

This set of simulations mirror those included in the main results but with only 10,000 individuals in each city. We can see increased variance between simulation runs as expected. Nonetheless, there is good concordance between the theoretical and simulation-based dominant eigenvalues and normalised eigenvalue. The dominant eigenvectors show more numerical instability, but the simulation-based results are still close to the theoretical eigenvectors with the same qualitative trends present.

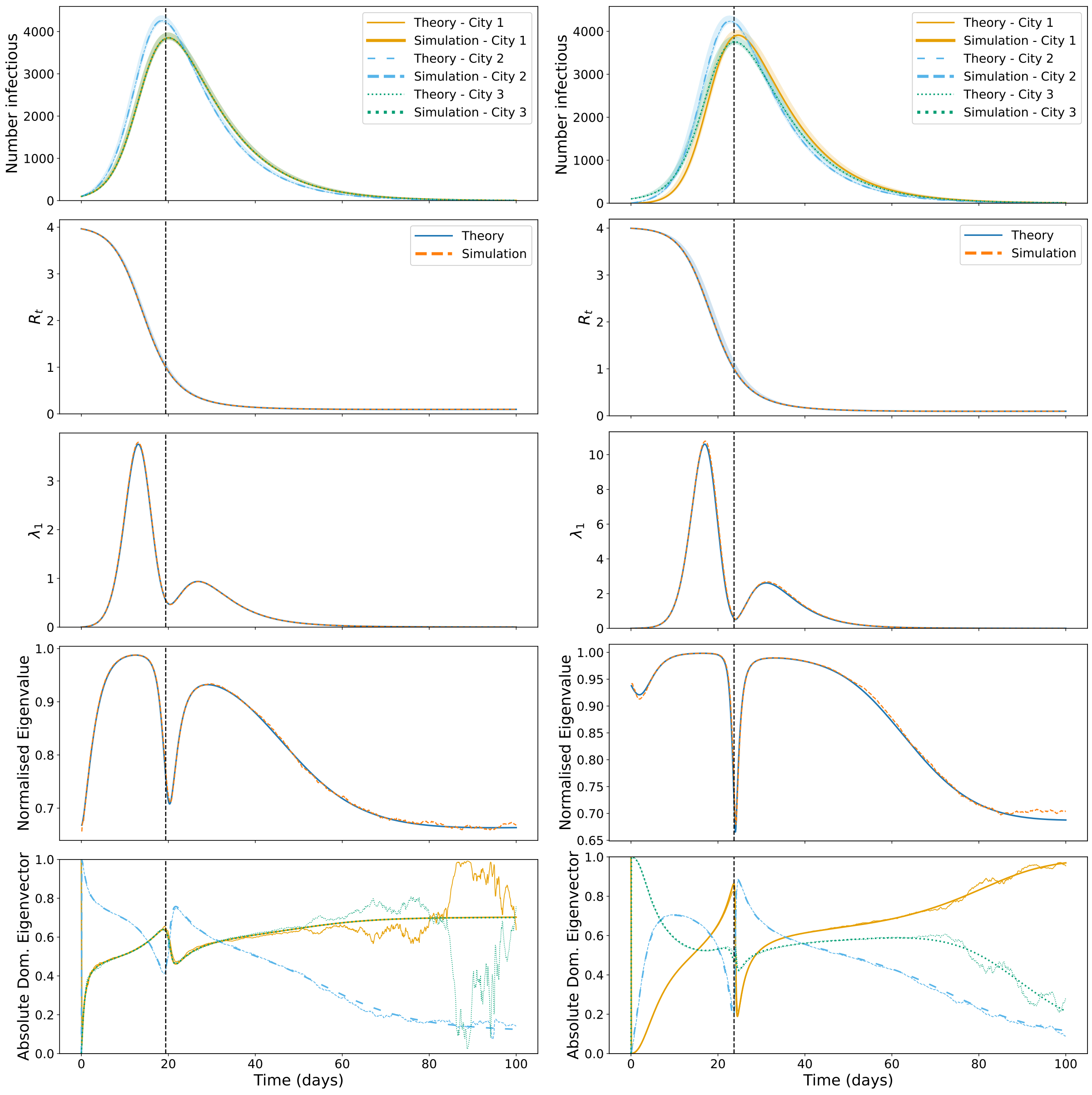


**(a)** Default infection scenario. **(b)** Seeding infection only in City 3.

**Figure S1. Outbreak dynamics and associated metrics. Top row** Infectious individuals in each city. **Second row** $R_t$ across the time period. **Third row** Dominant eigenvalue of the covariance matrix of the three cities. **Fourth row** $\lambda_1/\sqrt{\sum_i \lambda_i^2}$. **Bottom row** Absolute dominant eigenvector. Simulation results correspond to 100,000 realisations of the stochastic model with the mean plotted and the 95% prediction interval shown by the shaded area. **Left** Default infection scenario results. **Right** Results from seeding infection in City 3 only. The dashed vertical lines represent the epidemic transitions.

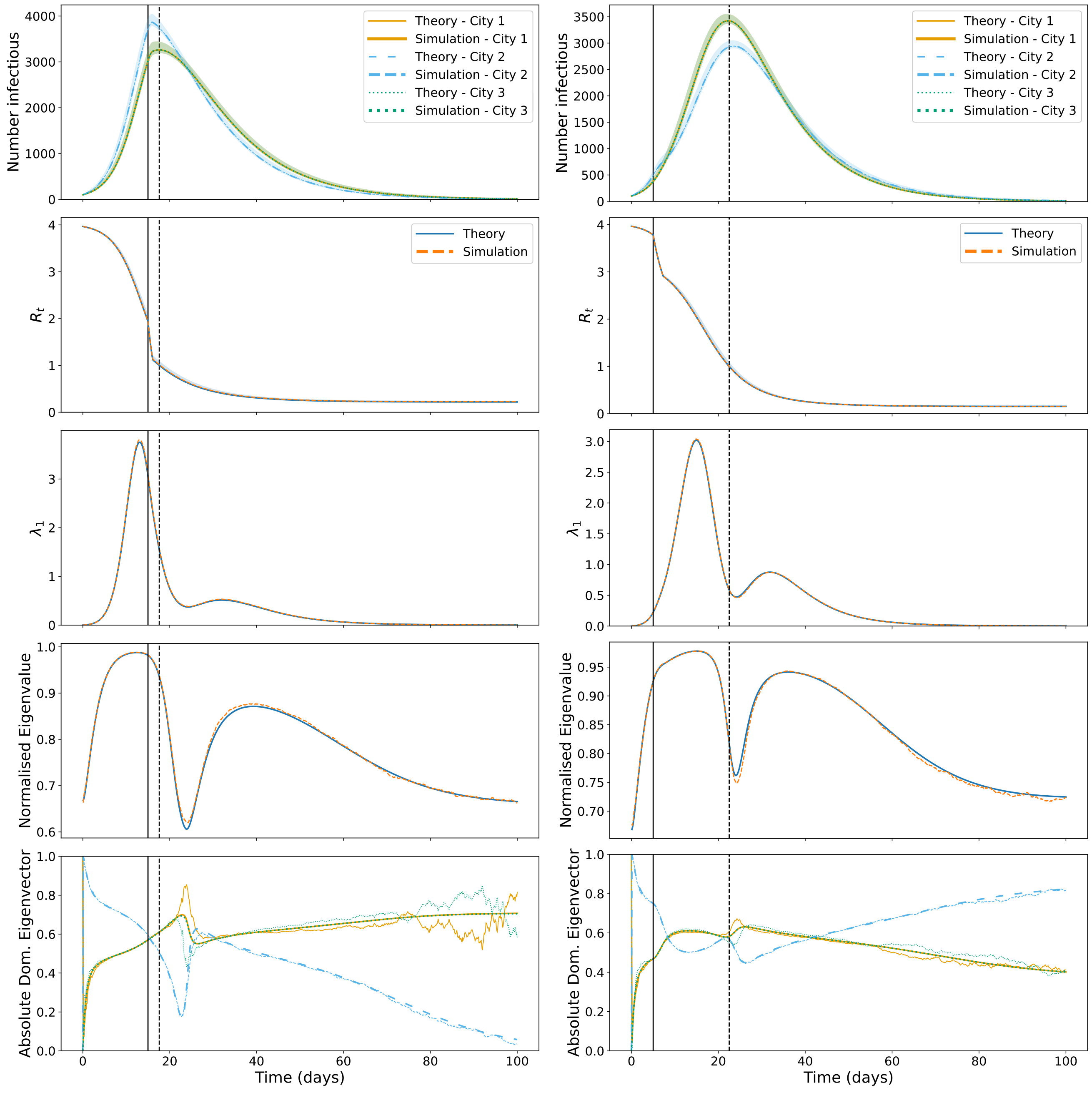


**(a)** Reducing transmission between cities.

**(b)** Reducing transmission only in City 2.

**Figure S2. Outbreak dynamics and associated metrics. Top row** Infectious individuals in each city. **Second row** $R_t$ across the time period. **Third row** Dominant eigenvalue of the covariance matrix of the three cities. **Fourth row** $\lambda_1/\sqrt{\sum_i \lambda_i^2}$. **Bottom row** Absolute dominant eigenvector. Simulation results correspond to 100,000 realisations of the stochastic model with the mean plotted and the 95% prediction interval shown by the shaded area. **Left** Results from reducing transmission between cities. **Right** Results from reducing transmission in City 2 only. The solid vertical lines indicate where transmission reduction begins and the dashed vertical lines represent the epidemic transitions.

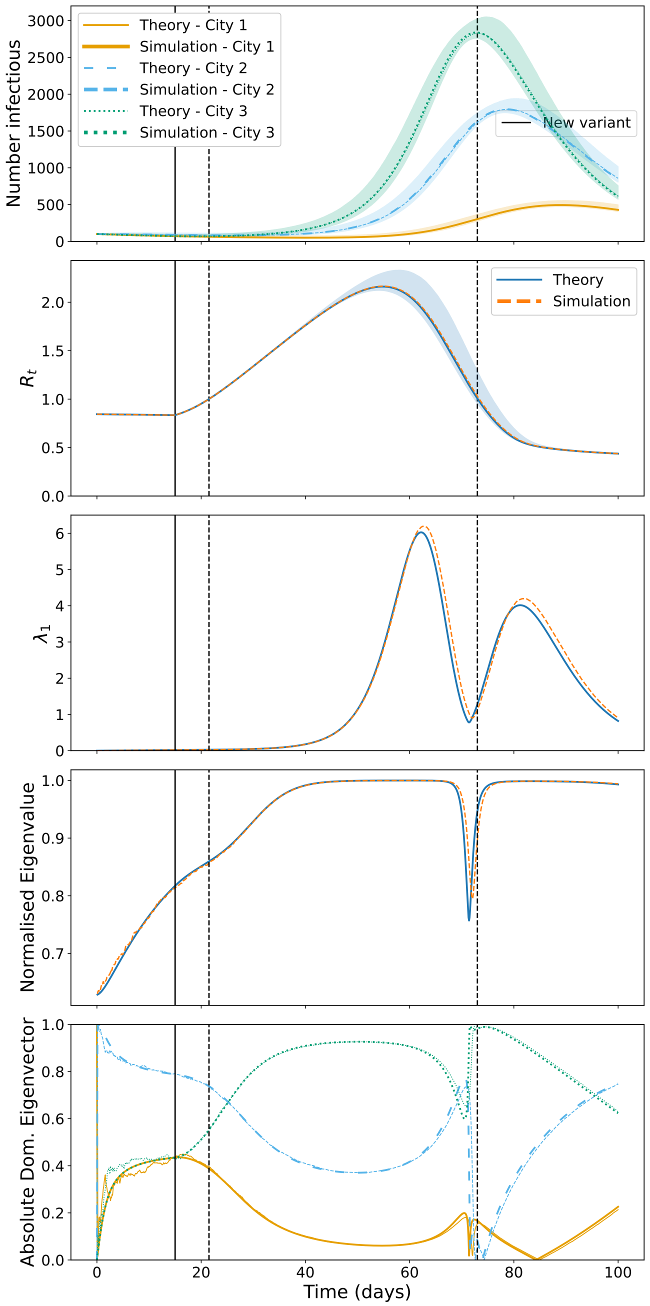


**Figure S3. Outbreak dynamics and associated metrics. Top row** Infectious individuals in each city. **Second row** $R_t$ across the time period. **Third row** Dominant eigenvalue of the covariance matrix of the three cities. **Fourth row** $\lambda_1/\sqrt{\sum_i \lambda_i^2}$. **Bottom row** Absolute dominant eigenvector. Simulation results correspond to 100,000 realisations of the stochastic model with the mean plotted and the 95% prediction interval shown by the shaded area. The solid vertical lines indicate the introduction of the new variant and the dashed vertical lines represent the epidemic transitions.

##### S1.2 Simulation incidence results

This set of results use the same simulations as in the main results but use the incidence of cases taken as the number of $I \to R$ transitions each day. Similar results are seen for the covariance matrix calculated using the daily incidence, but different trends are identified when the cumulative cases are used instead.

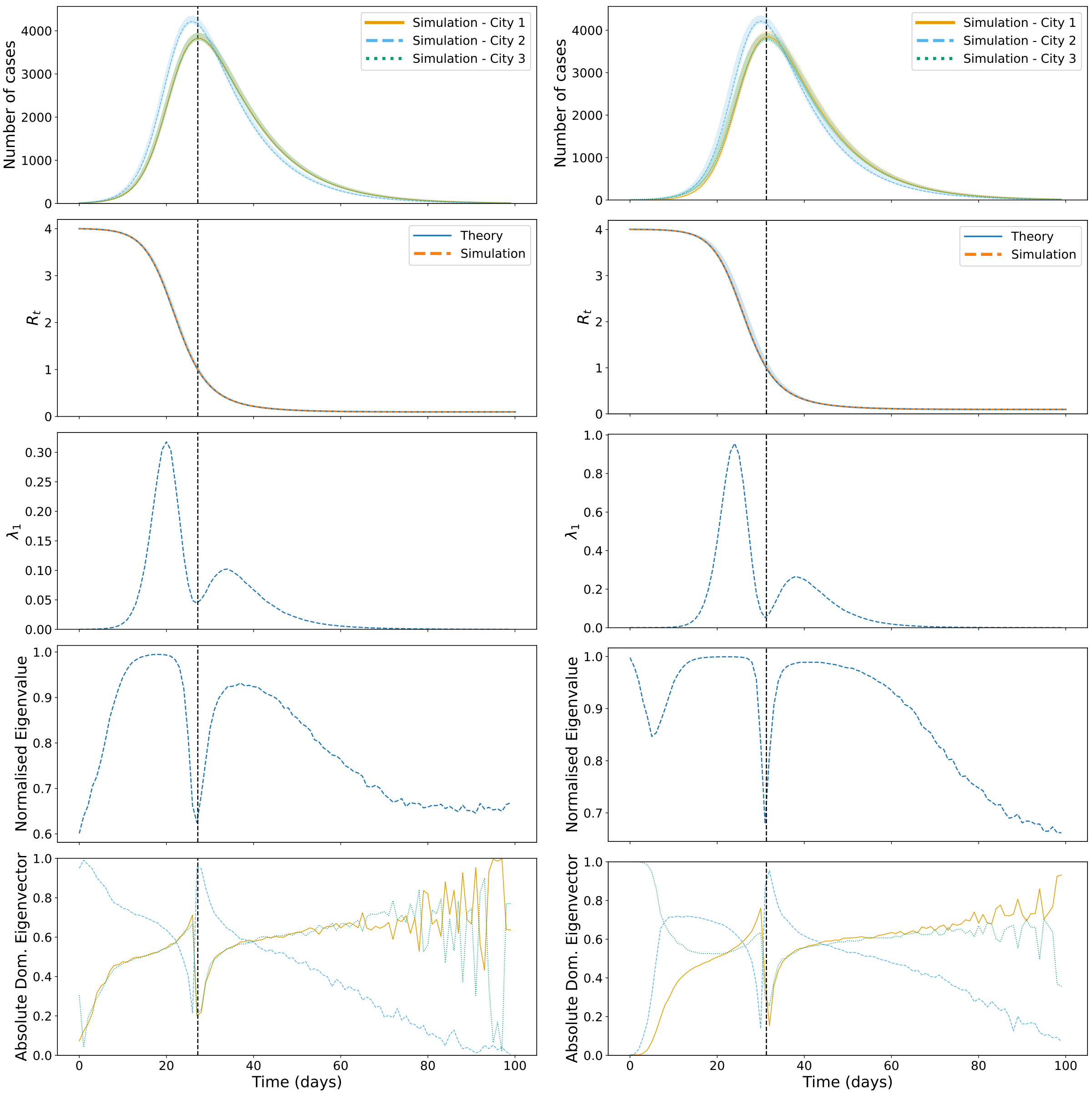


**(a)** Default infection scenario. **(b)** Seeding infection only in City 3.

**Figure S4. Outbreak dynamics and associated metrics. Top row** Number of cases in each city. **Second row** $R_t$ across the time period. **Third row** Dominant eigenvalue of the covariance matrix of the three cities. **Fourth row** $\lambda_1/\sqrt{\sum_i \lambda_i^2}$. **Bottom row** Absolute dominant eigenvector. Simulation results correspond to 100,000 realisations of the stochastic model with the mean plotted and the 95% prediction interval shown by the shaded area. **Left** Default infection scenario results. **Right** Results from seeding infection in City 3 only. The dashed vertical lines represent the epidemic transitions.

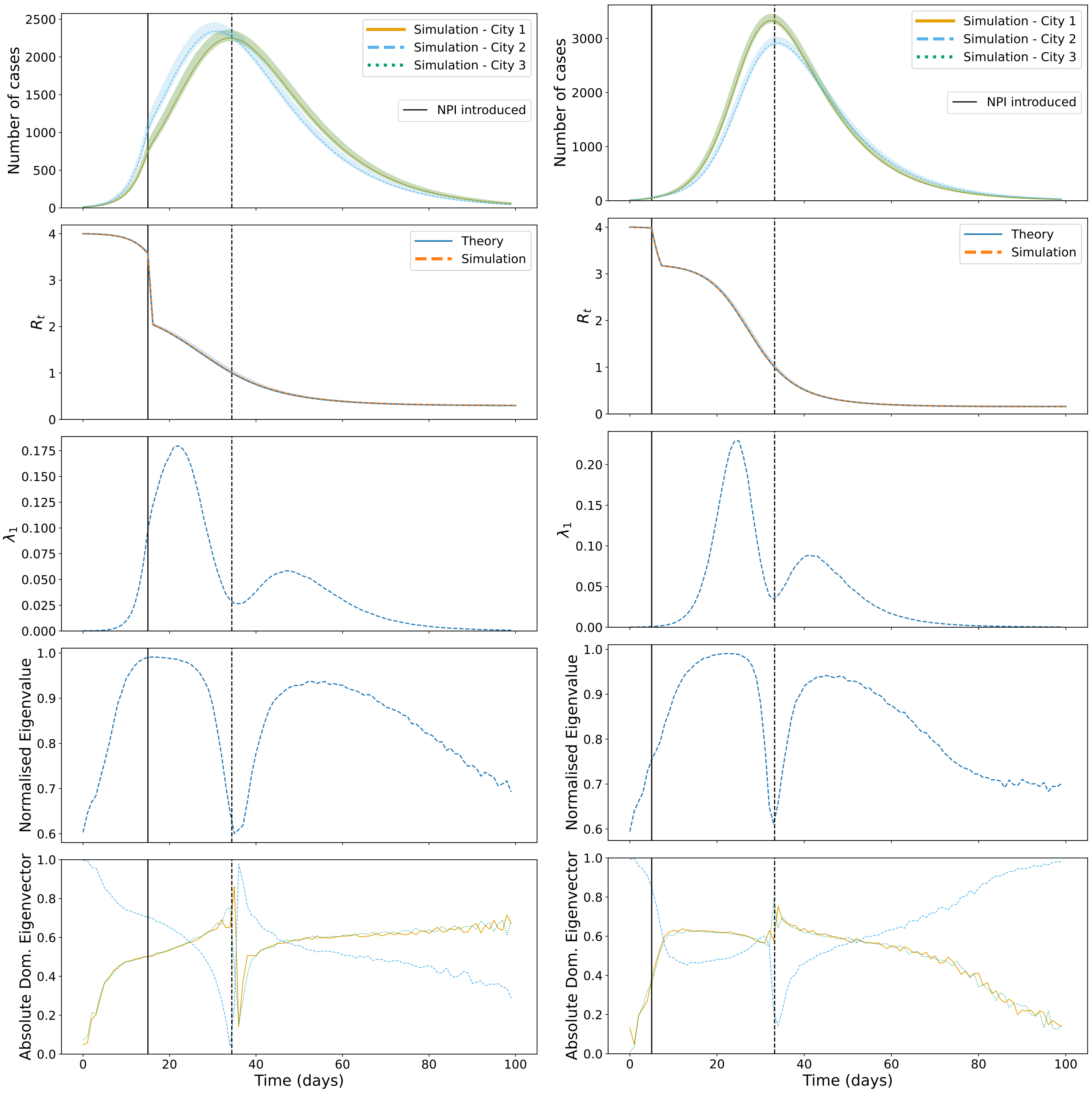


**(a)** Reducing transmission between cities. **(b)** Reducing transmission only in City 2.

**Figure S5. Outbreak dynamics and associated metrics. Top row** Number of cases in each city. **Second row** $R_t$ across the time period. **Third row** Dominant eigenvalue of the covariance matrix of the three cities. **Fourth row** $\lambda_1/\sqrt{\sum_i \lambda_i^2}$. **Bottom row** Absolute dominant eigenvector. Simulation results correspond to 100,000 realisations of the stochastic model with the mean plotted and the 95% prediction interval shown by the shaded area. **Left** Results from reducing transmission between cities. **Right** Results from reducing transmission in City 2 only. The solid vertical lines indicate where transmission reduction begins and the dashed vertical lines represent the epidemic transitions.

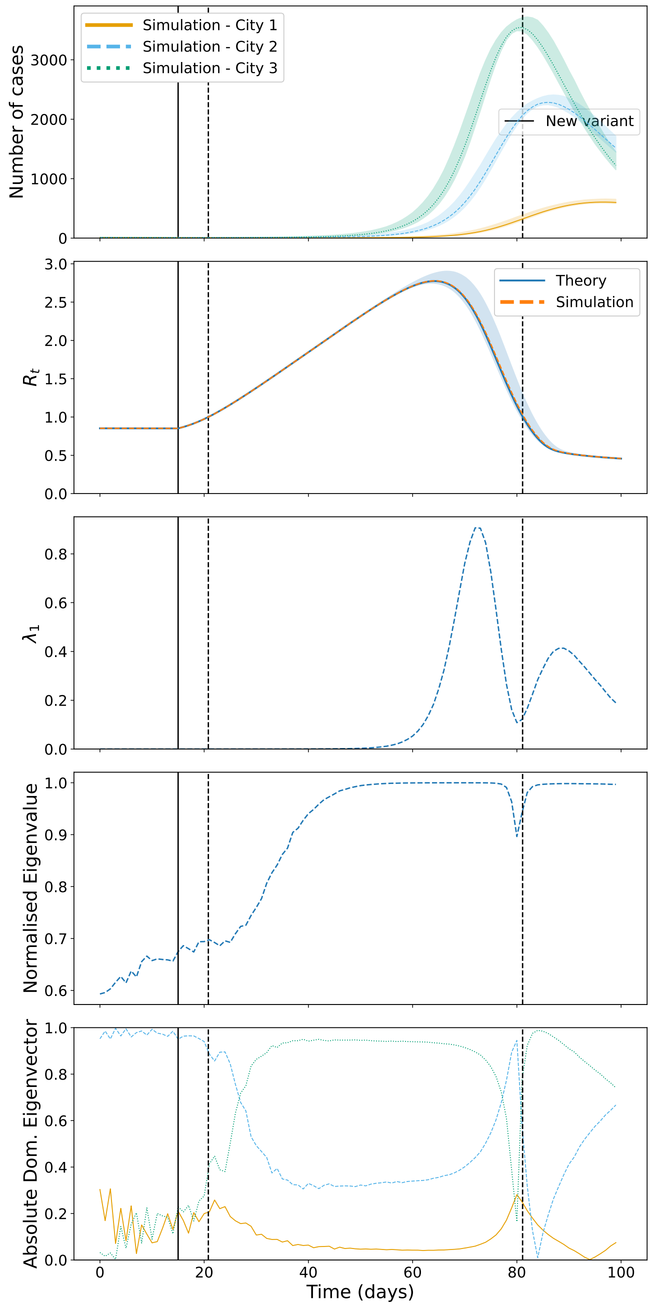


**Figure S6. Outbreak dynamics and associated metrics. Top row** Number of cases in each city. **Second row** $R_t$ across the time period. **Third row** Dominant eigenvalue of the covariance matrix of the three cities. **Fourth row** $\lambda_1/\sqrt{\sum_i \lambda_i^2}$. **Bottom row** Absolute dominant eigenvector. Simulation results correspond to 100,000 realisations of the stochastic model with the mean plotted and the 95% prediction interval shown by the shaded area. The solid vertical lines indicate the introduction of the new variant and the dashed vertical lines represent the epidemic transitions.

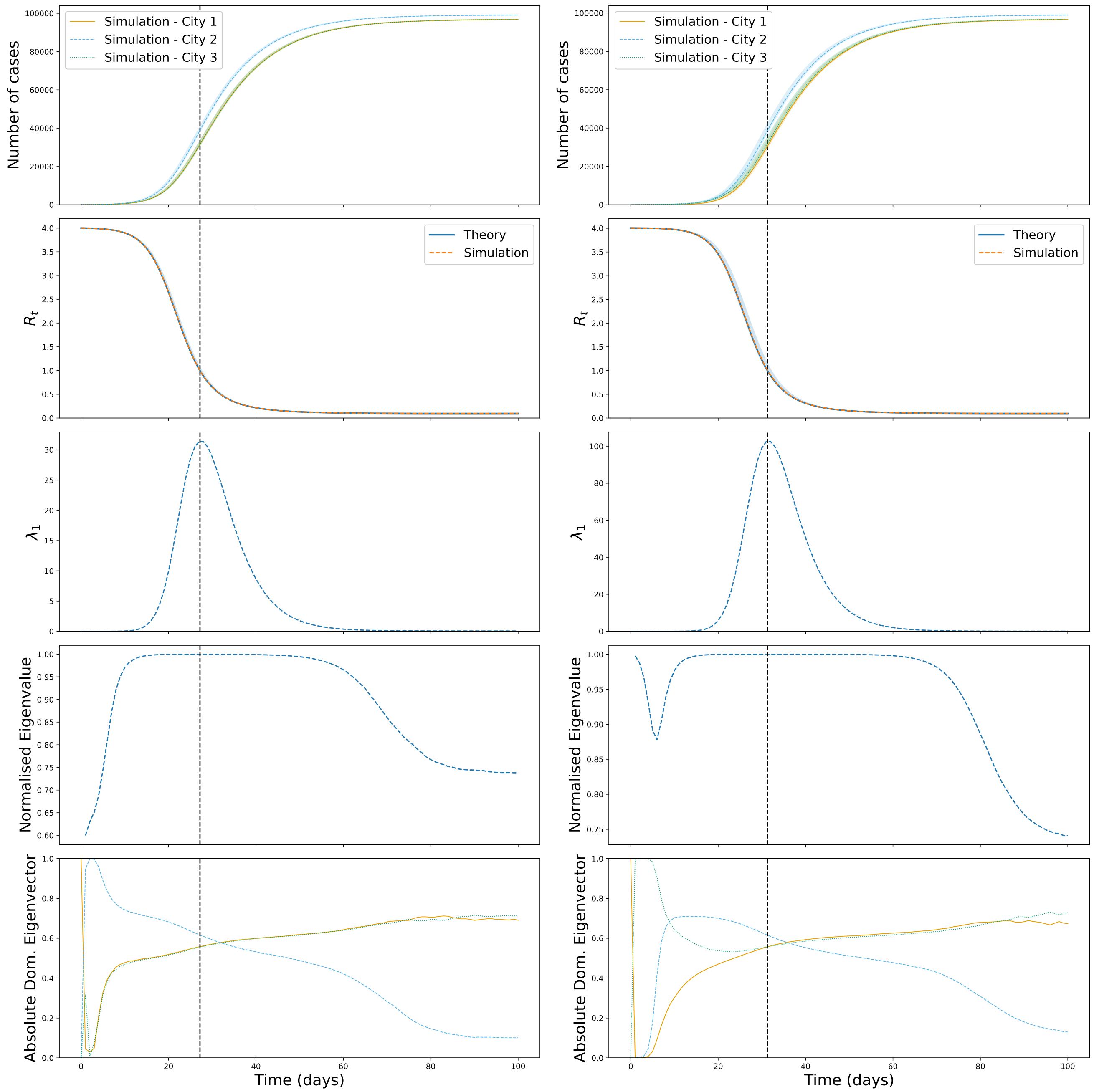


**(a)** Default infection scenario.

**(b)** Seeding infection only in City 3.

**Figure S7. Outbreak dynamics and associated metrics. Top row** Number of cumulative cases in each city. **Second row** $R_t$ across the time period. **Third row** Dominant eigenvalue of the covariance matrix of the three cities. **Fourth row** $\lambda_1/\sqrt{\sum_i \lambda_i^2}$. **Bottom row** Absolute dominant eigenvector. Simulation results correspond to 100,000 realisations of the stochastic model with the mean plotted and the 95% prediction interval shown by the shaded area. **Left** Default infection scenario results. **Right** Results from seeding infection in City 3 only. The dashed vertical lines represent the epidemic transitions.

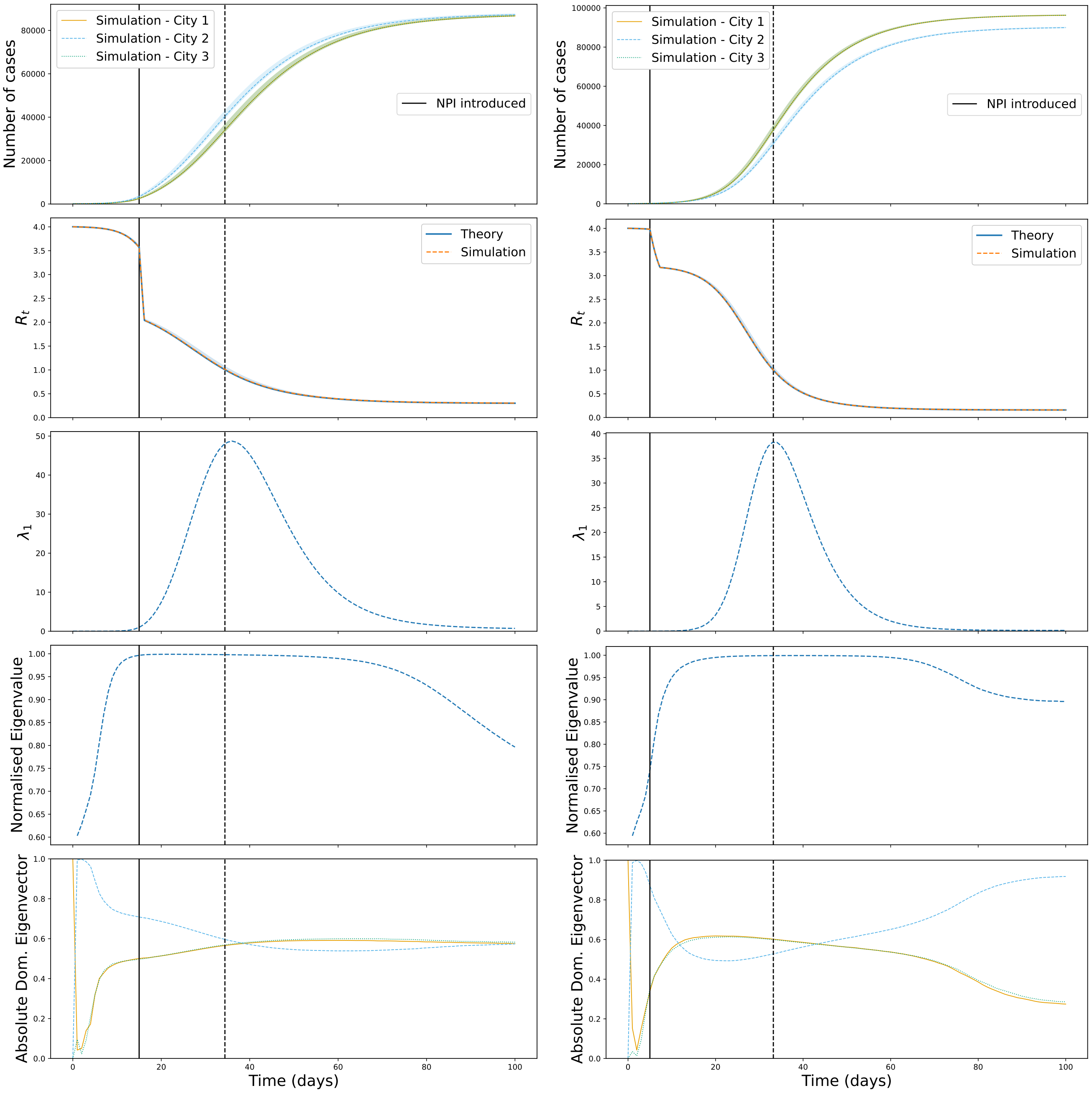


**(a)** Reducing transmission between cities. **(b)** Reducing transmission only in City 2.

**Figure S8. Outbreak dynamics and associated metrics. Top row** Number of cumulative cases in each city. **Second row** $R_t$ across the time period. **Third row** Dominant eigenvalue of the covariance matrix of the three cities. **Fourth row** $\lambda_1/\sqrt{\sum_i \lambda_i^2}$. **Bottom row** Absolute dominant eigenvector. Simulation results correspond to 100,000 realisations of the stochastic model with the mean plotted and the 95% prediction interval shown by the shaded area. **Left** Results from reducing transmission between cities. **Right** Results from reducing transmission in City 2 only. The solid vertical lines indicate where transmission reduction begins and the dashed vertical lines represent the epidemic transitions.

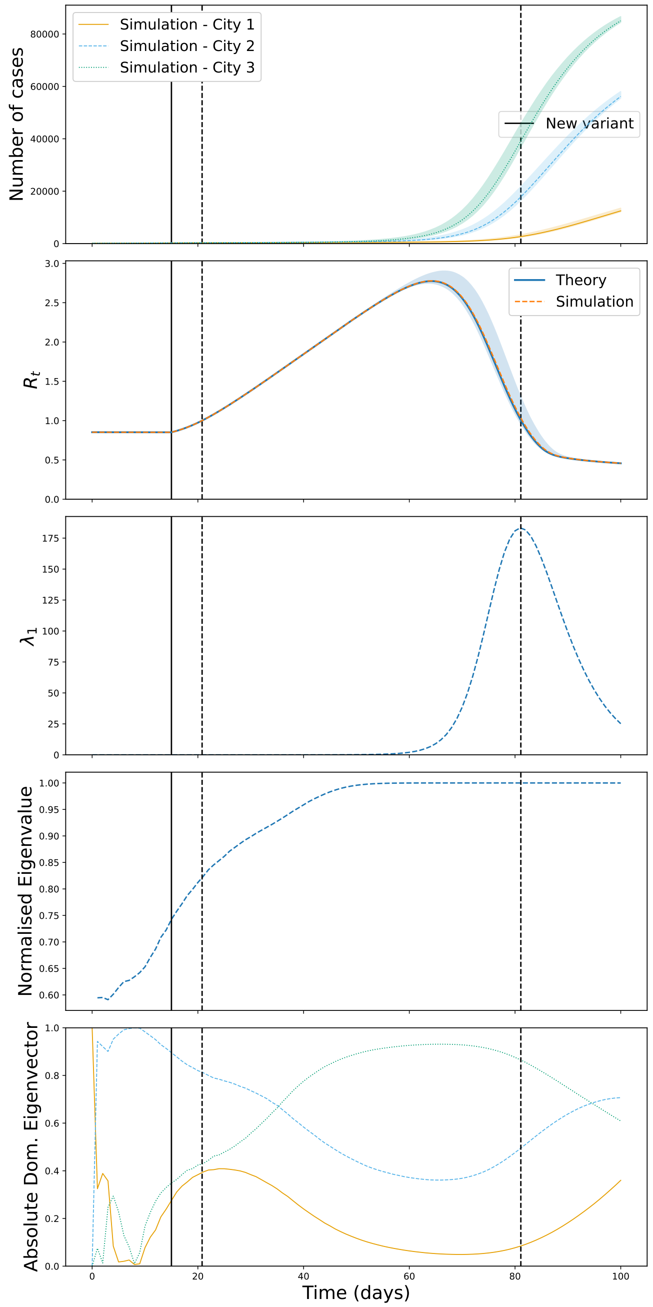


**Figure S9. Outbreak dynamics and associated metrics. Top row** Number of cumulative cases in each city. **Second row** $R_t$ across the time period. **Third row** Dominant eigenvalue of the covariance matrix of the three cities. **Fourth row** $\lambda_1/\sqrt{\sum_i \lambda_i^2}$. **Bottom row** Absolute dominant eigenvector. Simulation results correspond to 100,000 realisations of the stochastic model with the mean plotted and the 95% prediction interval shown by the shaded area. The solid vertical lines indicate the introduction of the new variant and the dashed vertical lines represent the epidemic transitions.

#### S1.3 5-year age-classes

We also considered the impacts of data aggregation on the calculated covariance matrix and eigendecomposition. In Fig. S10, the eigenvalue-based early warning signals behave in almost exactly the same manner as in the 10-year aggregated results, apart from the dominant eigenvalue being approximately double the size (due to the halving of the age-bands).

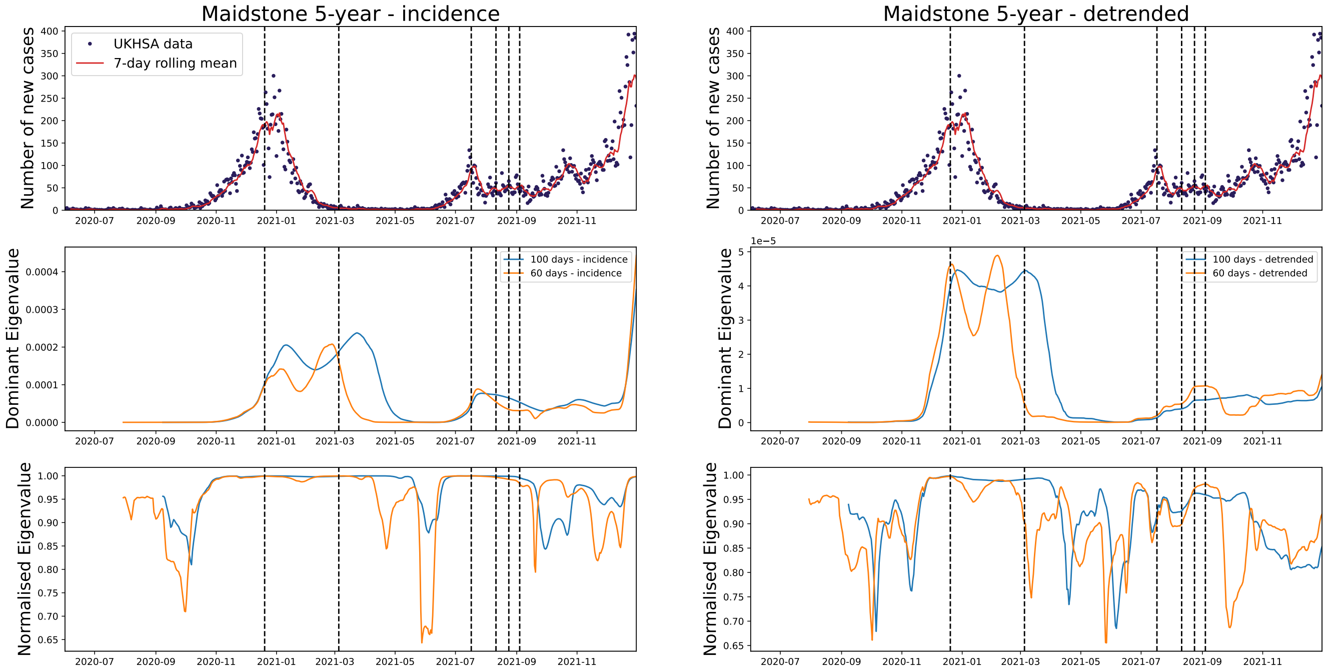


**Figure S10. Top** Incidence and rolling mean of COVID-19 cases in Maidstone. **Middle** Dominant eigenvalue of the covariance matrix of the 5-year age-aggregated incidence (**left**) and detrended (**right**) time series using a rolling window of 100 and 60 days. **Bottom** $\lambda_1/\sqrt{\sum_i \lambda_i^2}$.

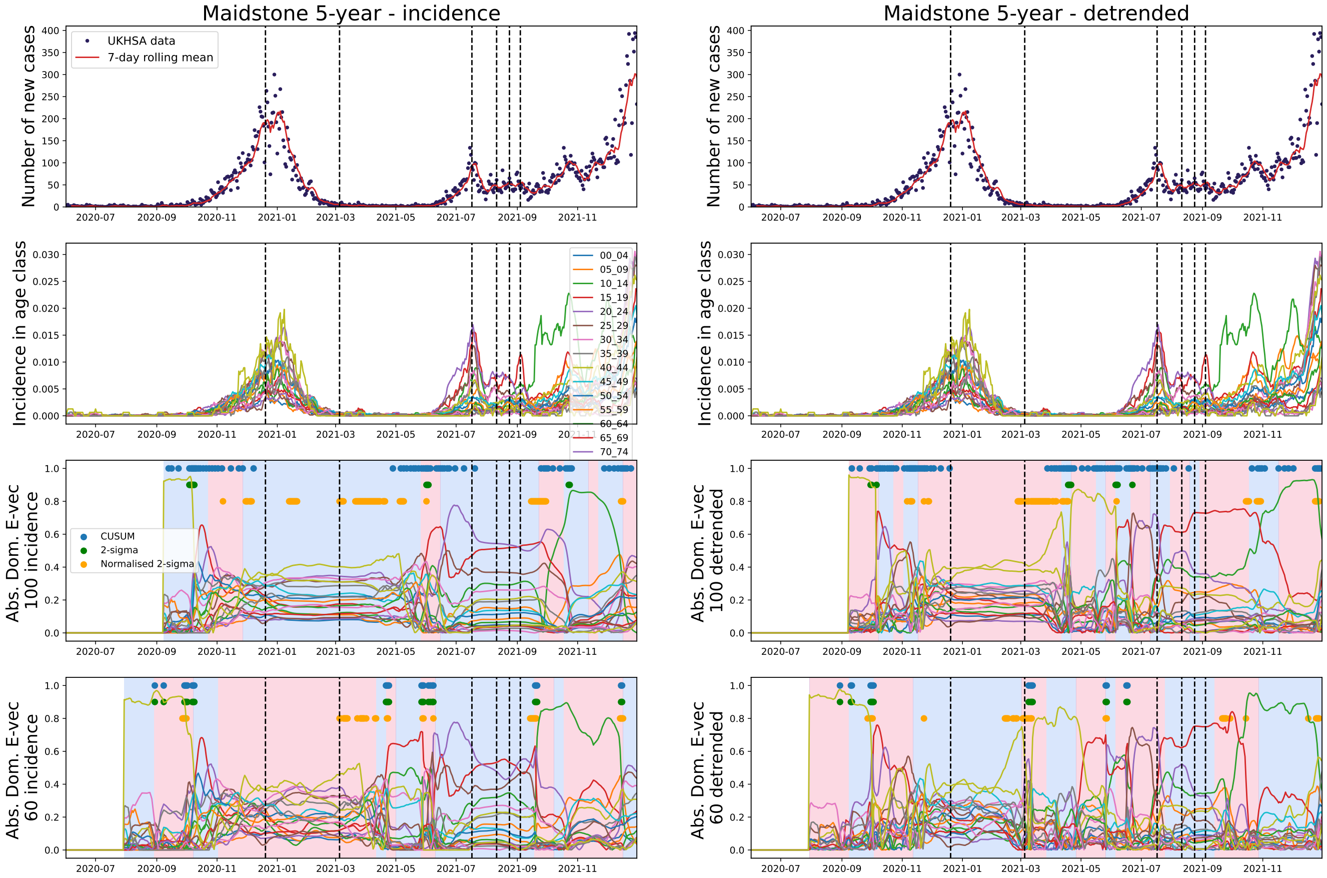


**Figure S11. Top row** Incidence and rolling mean of COVID-19 cases in Maidstone. **Second row** Incidence time series for each age-class. **Third row** Absolute value of the dominant eigenvector of the covariance matrix of the 5-year age-aggregated incidence (**left**) and detrended (**right**) time series using a rolling window of 100 days. **Bottom** Absolute value of the dominant eigenvector of the covariance matrix of the 5-year age-aggregated incidence (**left**) and detrended (**right**) time series using a rolling window of 60 days. Dashed vertical lines show epidemic transitions, solid dots indicate where each method signals a change in the angular rotation and background colour changes are change-points identified by linearly penalised segmentation.

#### S1.4 30-day rolling windows

We conducted a sensitivity analysis into the rolling window size used when estimating the covariance matrix of the age-structured data. Figs. S12 and S13 show that similar trends are seen as when using 100-day or 60-day rolling windows, but the results are much more stochastic.

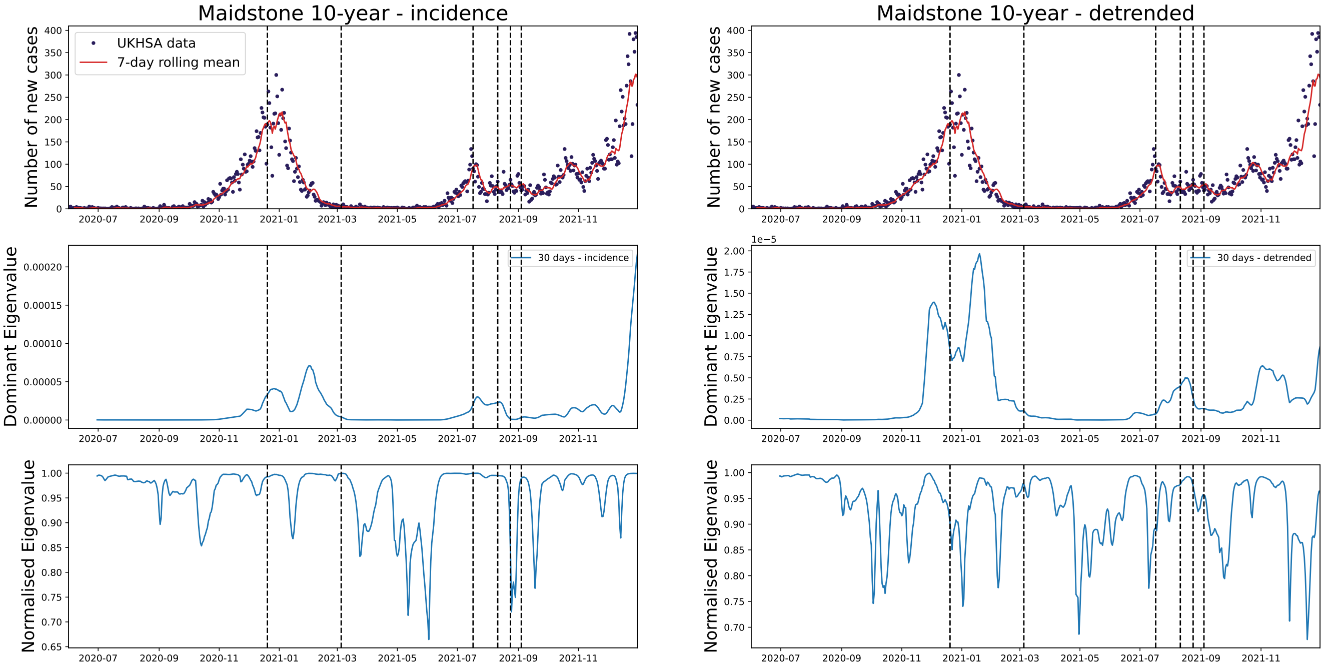


**Figure S12. Top** Incidence and rolling mean of COVID-19 cases in Maidstone. **Middle** Dominant eigenvalue of the covariance matrix of the 10-year age-aggregated incidence (**left**) and detrended (**right**) time series using a rolling window of 30 days. **Bottom** $\lambda_1/\sqrt{\sum_i \lambda_i^2}$.

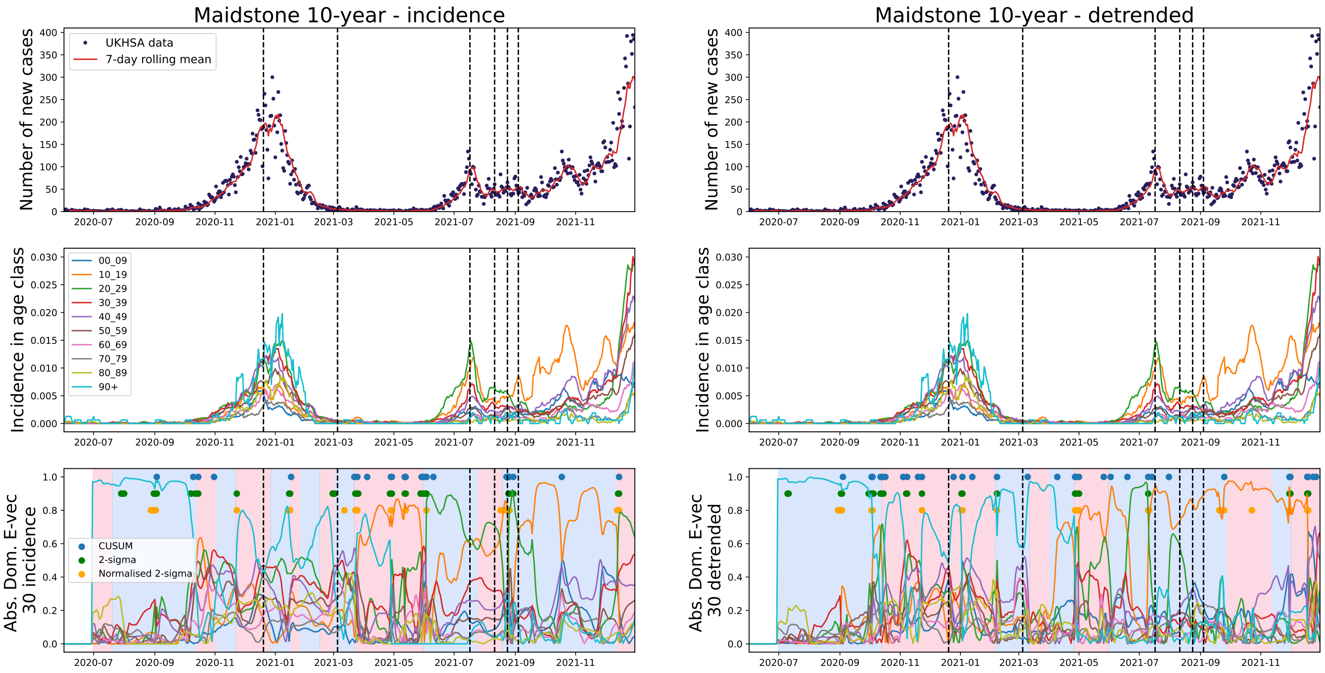


**Figure S13. Top row** Incidence and rolling mean of COVID-19 cases in Maidstone. **Second row** Incidence time series for each age-class. **Third row** Absolute value of the dominant eigenvector of the covariance matrix of the 10-year age-aggregated incidence (**left**) and detrended (**right**) time series using a rolling window of 30 days. Dashed vertical lines show epidemic transitions, solid dots indicate where each method signals a change in the angular rotation and background colour changes are change-points identified by linearly penalised segmentation.

#### S2 Metapopulation model matrices

Here we return to the simple model metapopulation model and give the explicit forms of the Jacobian and diffusion matrices. These are block matrices with the following forms:

$$J = \begin{bmatrix} J_{SS} & J_{SI} \\ J_{IS} & J_{II} \end{bmatrix}, \quad B = \begin{bmatrix} B_{SS} & B_{SI} \\ B_{IS} & B_{II} \end{bmatrix}, \tag{S1}$$

where the $3 \times 3$ submatrices are:

$$J_{SS} = \operatorname{diag}(-\mu - \sum_{k=1}^{3} \beta_{1k} i_k, -\mu - \sum_{k=1}^{3} \beta_{2k} i_k, -\mu - \sum_{k=1}^{3} \beta_{3k} i_k) \tag{S2}$$

$$J_{SI\,jk} = -\beta_{jk} s_j \tag{S3}$$

$$J_{IS} = \operatorname{diag}(\sum_{k=1}^{3} \beta_{1k} i_k, \sum_{k=1}^{3} \beta_{2k} i_k, \sum_{k=1}^{3} \beta_{1k} i_k, \sum_{k=1}^{3} \beta_{1k} i_k) \tag{S4}$$

$$J_{II\,jk} = -(\mu + \gamma)\delta_{jk} + \beta_{jk} s_j \tag{S5}$$

$$B_{SS} = \operatorname{diag}(\mu(1 - s_1) + \sum_{k=1}^{3} \beta_{1k} s_1 i_k, \mu(1 - s_2) + \sum_{k=1}^{3} \beta_{2k} s_2 i_k, \mu(1 - s_3) + \sum_{k=1}^{3} \beta_{3k} s_3 i_k) \tag{S6}$$

$$B_{SI} = B_{IS} = \operatorname{diag}(-\mu i_1 - \sum_{k=1}^{3} \beta_{1k} s_1 i_k, -\mu i_2 - \sum_{k=1}^{3} \beta_{2k} s_2 i_k, -\mu i_3 - \sum_{k=1}^{3} \beta_{3k} s_3 i_k) \tag{S7}$$

$$B_{II} = \operatorname{diag}((\gamma + \mu) i_1 + \sum_{k=1}^{3} \beta_{1k} s_1 i_k, (\gamma + \mu) i_2 + \sum_{k=1}^{3} \beta_{2k} s_1 i_k, (\gamma + \mu) i_3 + \sum_{k=1}^{3} \beta_{3k} s_3 i_k) \tag{S8}$$

and $\delta_{jk}$ is the Kronecker-delta function.

#### S3 Eigenvalue Scaling of Aggregated Cases

In this section, we explore how aggegration affects the eigenvalues of the covariance matrix, especially with respect to the patterns seen between the 5-year and 10-year age-class results. Specifically, we consider a system of $1,2,\ldots,n$ age-classes each with $N_i$ individuals and then the aggregated system with $1,2,\ldots,n/2$ age-classes each with $N_1+N_2,\ldots,N_{n/2-1}+N_{n/2}$ individuals. Taking a noise-scaling given by the system-size expansion and using the linear noise approximation, we can then prove the following relationship between the eigenvalues of the aggregated and dis-aggregated covariance matrices:

**Proposition 1.** *Let $X=(x_1,x_2,\ldots,x_n)\sim N(0,\Sigma)$ be a set of stochastic variables given by a multivariate Gaussian distribution with mean 0 and covariance $\Sigma$ (such as those derived from the linear noise approximation). Let $Y=(y_1,y_2,\ldots,y_{n/2})\sim N(0,\Gamma)$ be defined similarly, where each $y_i=\frac{N_{2i-1}^{1/2}x_{2i-1}+N_{2i}^{1/2}x_{2i}}{\sqrt{M_i}}$, $\quad M_i=N_{2i}+N_{2i-1}$. Let $\lambda_1',\lambda_2',\ldots,\lambda_{n/2}'$ be the eigenvalues of $\Gamma$ in descending order and $\lambda_1,\lambda_2,\ldots,\lambda_n$ be the eigenvalues of $\Sigma$ arranged similarly.*
*Then $\lambda_{1+n/2}\leq\lambda_1'\leq\lambda_1$.*

*Proof.* We note that $Y=WX$, where

$$W_{ij}=\begin{cases}\sqrt{\frac{N_{2i-1}}{M_i}} & j=2i-1,\\ \sqrt{\frac{N_{2i}}{M_i}} & j=2i,\\ 0 & \text{otherwise,}\end{cases}$$

and $W$ is an $\mathbb{R}^{n/2\times n}$ semi-orthogonal matrix such that $WW^T=I_{n/2}$, where $I_{n/2}$ is the identity matrix of size $n/2$. Further, $\Gamma,\Sigma$ are real, symmetric matrices (as they are covariance matrices) and then
$\Gamma=Cov(Y)=\mathbb{E}(YY^T)=\mathbb{E}(WXX^TW^T)=W\mathbb{E}(XX^T)W^T=W\Sigma W^T$. Then the relation follows directly from the Poincaré separation theorem (Cauchy interlacing theorem) for orthogonal projections. □

**Remark 1.1.** *Because the covariance operator is translation-invariant (i.e.* $\mathrm{Var}(X+c)=\mathrm{Var}(X)$*), this eigenvalue relationship holds regardless of the means of $X$ and $Y$.*

In the case of the stochastic residuals in the 5-year age-class and 10-year age-class results, this indicates that the dominant eigenvalue of the 10-year age class is bounded above by the dominant eigenvalue of the 5-year age-class.
Of interest to infectious disease modellers is the impact on observed statistics due to aggregation/disaggregation. We can also derive a lower bound on the aggregated dominant eigenvalue as a function of the disaggregated variances and covariances. In lower-dimensional systems, an increase in variance is a known EWS of a change in Jacobian spectrum/criticality and so we aim to express our dominant eigenvalue by these increasing statistics.

**Proposition 2.** *Let $Y=WX$ with $X,Y$ defined as above. Then the dominant eigenvalue of the aggregated system, $\lambda_1'$, satisfies the inequality*

$$\lambda_1'\geq\max_i\Big(\frac{1}{\sum_j W_{ij}^2}\sum_j W_{ij}^2 Var(x_j)+\sum_{k\neq j}(W_{ij}W_{ij}Cov(x_k,x_j)\Big).$$

*Proof.* We first note that because $\Gamma$ is a positive semi-definite matrix, by the Rayleigh quotient we have that $\lambda_1'\geq\max_i\Gamma_{ii}=\max_i\mathrm{Var}(y_i)$. Thus, expanding the variance we have

$$\lambda_1'\geq\max_i\Big(\frac{1}{\sum_j W_{ij}^2}\sum_j W_{ij}^2 Var(x_j)+\sum_{k\neq j}(W_{ij}W_{ij}Cov(x_k,x_j)\Big).$$

□

Returning to the case under the linear noise approximation, this means that $\lambda_1$ is bounded below by a function of the covariances between each aggregated pair of $x_i$:

$$\lambda_1'\geq\max_i\Big(\frac{1}{M_i}[N_{2i-1}Var(x_{2i-1})+N_{2i}Var(x_{2i})+2\sqrt{N_{2i-1}N_{2i}}Cov(x_{2i-1},x_{2i})]\Big)$$

Intuitively, the variance of each $x_i$ increases due to critical slowing down where the only negative term on the right hand side may be the covariances between compartments (which are likely positive for infectious residuals as residuals become more correlated closer to a change in Jacobian spectrality and due to infections). In terms of EWSs, although being bounded by increasing functions does not guarantee $\lambda_1'$ increases, combined with the expectation $\lambda_1'$ that is of the same magnitude to its bounds it does suggest that changes in $\lambda_1'$ hold predictive power. Further, it also indicates that oscillations in $\lambda_1'$ depend on the differences in the rate of increase in these bounds.

Beyond the dominant eigenvalue, we can quantify the exact loss in total variance (and therefore stochasticity) from aggregation.

**Proposition 3.** *Let $X$ and $Y = WX$ be defined as above. The total variance of the disaggregated system is* $\text{tr}(\Sigma) = \sum_{i=1}^{n} \lambda_i$, *where* tr *is the trace of the matrix, and the total variance of the aggregated system is* $\text{tr}(\Gamma) = \sum_{i=1}^{n/2} \lambda_i'$. *Then the following inequality holds:*

$$\text{tr}(\Sigma) - \text{tr}(\Gamma) \geq 0$$

*Proof.* We note that this holds immediately from the proof of the previous proposition on the bounds of the eigenvalues but seek a geometric reasoning to show that the loss of information is to the projection operator. Using the cyclic property of the trace, we can write $\text{tr}(\Gamma) = \text{tr}(W^T \Sigma W) = \text{tr}(W^T W \Sigma)$. We can then express the difference in total variance as:

$$\text{tr}(\Sigma) - \text{tr}(\Gamma) = \text{tr}(\Sigma) - \text{tr}(W^T W \Sigma) = \text{tr}((I_n - W^T W)\Sigma).$$

Because $W$ is semi-orthogonal, $W^T W$ is an orthogonal projection matrix. Consequently, $I - W^T W$ is the complementary projection matrix onto the null space of $W$. As it is also a projection matrix it is positive semi-definite. Since $\Sigma$ is strictly positive definite, the trace of their product is non-negative, proving that $\text{tr}(\Gamma) \leq \text{tr}(\Sigma)$. □

This shows that the variance loss is due to the projection of high-dimensional dynamics (the disaggregated data) into a lower-dimensional subspace.

#### S4 Non-stationary Covariance Evolution

This appendix contains the mathematical derivations of the main EWSs tested in the paper, namely the behaviour of the dominant eigenvalue and eigenvector of the covariance matrix.

##### S4.1 Assumptions and Hypothesis

We consider the Fokker-Planck Equation for the probability distribution $\Pi(\boldsymbol{\xi},t)$ of a set of $N$ variables $\xi_1,\dots,\xi_N$ in the usual form:

$$\frac{\mathrm{d}\Pi(\boldsymbol{\xi},t)}{\mathrm{d}t} = -\sum_i \frac{\partial}{\partial \xi_i}\left(\sum_j J_{i,j}\xi_j\Pi\right) + \frac{1}{2}\sum_i\sum_j B_{ij}\frac{\partial^2\Pi}{\partial\xi_i\partial\xi_j} \tag{S9}$$

where the stochastic variables represent the deviations from the deterministic solutions of a dynamical system with Jacobian $J$ and where $B$ is an intrinsic noise matrix derived from the system-size expansion. The Lyapunov equation for the covariance matrix $\Sigma$ is then:

$$\frac{\mathrm{d}\Sigma}{\mathrm{d}t} = J\Sigma + \Sigma J^T + B. \tag{S10}$$

We assume that $\Sigma$ is positive definite and has a non-degenerate spectrum with eigendecomposition: $\Sigma = Q\Lambda Q^T$, where $\Lambda$ is a diagonal matrix of eigenvalues and the columns of $Q$ are the orthonormal eigenvectors of the system. Note that $QQ^T = I$ where $I$ is the identity matrix

Then, we claim that the rate of rotation of the eigenbasis, $Q\frac{\mathrm{d}(Q^T)}{\mathrm{d}t}$, depends on the off-diagonal elements of the projection of $J$ and $B$ onto $Q$ (i.e. $(Q^TJQ)_{ij}$ and $(Q^TBQ)_{ij}$). We also expect it to increase on approach to a critical transition, in periods of non-stationarity and when the time series deviates the most from the deterministic solutions to the dynamic system. Thus, it should provide an EWS of epidemic transitions.

##### S4.2 (Near)-Stationary Eigenvector Evolution

We first consider the idealised scenario where the covariance matrix of the system can perfectly capture the changes in $J$ and $B$ such that the eigenbasis of $\Sigma$ is stationary. We note that $\frac{\mathrm{d}Q}{\mathrm{d}t} = 0$, such that $\Pi$ has reached its stationary Gaussian distribution, is a special case of this scenario and is usually assumed in the literature.
To begin, we prove the following statement:

**Proposition 4.** *If $J = Q\Lambda_J Q^T, B = Q\Lambda_B Q^T$ (i.e. $J$ and $B$ are diagonalisable by $Q$), then $\frac{\mathrm{d}Q}{\mathrm{d}t} = 0$.*

*Proof.* Using the eigendecomposition of $\Sigma$ the time-derivative of $\Sigma$ can then be expanded as:

$$\frac{\mathrm{d}\Sigma}{\mathrm{d}t} = \left(\frac{\mathrm{d}Q}{\mathrm{d}t}\right)\Lambda Q^T + Q\left(\frac{\mathrm{d}\Lambda}{\mathrm{d}t}\right)Q^T + Q\Lambda\left(\frac{\mathrm{d}(Q^T)}{\mathrm{d}t}\right) = J\Sigma + \Sigma J^T + B \tag{S11}$$

Pre- and post-multiplying by $Q^T, Q$ respectively and denoting $M = Q^T(\frac{\mathrm{d}Q}{\mathrm{d}t}) = -Q(\frac{\mathrm{d}(Q^T)}{\mathrm{d}t})$ gives:

$$M\Lambda - \Lambda M + \frac{\mathrm{d}\Lambda}{\mathrm{d}t} = Q^TJQ\Lambda + \Lambda Q^TJ^TQ + Q^TBQ \tag{S12}$$

and by our assumption on the diagonalisability of $J$ and $B$ by $Q$, we know that the only possibly non-diagonal matrix is $M\Lambda - \Lambda M$. However, as the RHS is the sum of diagonal matrices, this also means that $A_{ij} = (M\Lambda - \Lambda M)_{ij} = 0$ and

$$(M\Lambda - \Lambda M)_{ij} = M_{ij}(\lambda_j - \lambda_i) \tag{S13}$$

where we also note that $A_{ii} = 0$ as it is an antisymmetric matrix. For the off-diagonal elements of $A$ to be 0, either $\Sigma$ has a degenerate spectrum or $M_{ij} = 0$. From our assumption that $\Sigma$ has a non-degenerate spectrum we can then conclude that $M = 0$. Finally, we note that:

$$Q(0) = Q(M) \tag{S14}$$

$$= Q\left(Q^T\frac{\mathrm{d}Q}{\mathrm{d}t}\right) = (QQ^T)\frac{\mathrm{d}Q}{\mathrm{d}t} = I\frac{\mathrm{d}Q}{\mathrm{d}t} \tag{S15}$$

$$\implies \frac{\mathrm{d}Q}{\mathrm{d}t} = 0 \tag{S16}$$

where the last line follows as $I \neq 0$ and so $Q$ is constant. □

Although this may be the case for certain $J$ and $B$ (such as when $B$ is a constant matrix), this is unlikely to hold true for a general dynamical system. Thus, we now provide a necessary and sufficient condition for $\frac{\mathrm{d}Q}{\mathrm{d}t} = 0$ in Proposition 5.

**Proposition 5.** $\frac{\mathrm{d}Q}{\mathrm{d}t} = 0 \iff \lambda_j(Q^TJQ)_{ij} + \lambda_i(Q^TJ^TQ)_{ij} + (Q^TBQ)_{ij} = 0 \forall i \neq j$

*Proof.* For the forward direction, we assume $\frac{\mathrm{d}Q}{\mathrm{d}t} = 0$ and return to the diagonalised Lyapunov equation for $\Sigma$:

$$M\Lambda - \Lambda M + \frac{\mathrm{d}\Lambda}{\mathrm{d}t} = Q^TJQ\Lambda + \Lambda Q^TJ^TQ + Q^TBQ \tag{S17}$$

$$\frac{\mathrm{d}\Lambda}{\mathrm{d}t} = Q^TJQ\Lambda + \Lambda Q^TJ^TQ + Q^TBQ \tag{S18}$$

$$\implies (Q^TJQ\Lambda + \Lambda Q^TJ^TQ + Q^TBQ)_{ij} = 0 \tag{S19}$$

where we note that the last line follows as $\frac{\mathrm{d}\Lambda}{\mathrm{d}t}$ is the time derivative of a diagonal matrix and so is also a diagonal matrix. Thus, the right hand side of the equation must also be a diagonal matrix.
For the reverse direction, the assumption that $\lambda_j(Q^TJQ)_{ij} + \lambda_i(Q^TJ^TQ)_{ij} + (Q^TBQ)_{ij} = 0$ again makes the right hand side of the Lyapunov equation a diagonal matrix. We know that the only possible off-diagonal terms that arise on the left hand side are in $M\Lambda - \Lambda M$, and so using identical logic to the proof of Proposition 4 we can conclude that $\frac{\mathrm{d}Q}{\mathrm{d}t} = 0$. ☐

Unfortunately, this condition is restrictive when considering real-world dynamical systems, especially if we are considering time-dependent $J$ and $B$. However, close to a stationary point, $Q^TJQ$ and $Q^TBQ$ are likely to have much smaller off-diagonal terms; $Q$ captures most of the relevant dynamics of $J$ and $B$ such that the change of basis matrix is relatively constant.

**Remark 5.1.** *If the system is assumed to be stationary, such that $J$ and $B$ are constant matrices and $\frac{\mathrm{d}\Sigma}{\mathrm{d}t} = 0$, then $\frac{\mathrm{d}Q}{\mathrm{d}t} = 0$, trivially.*

##### S4.3 Rate of Rotation of the Eigenbasis

As noted in the previous section, the restrictive conditions required for the proofs likely do not represent data from real-world disease systems, especially when considering the additional stochasticity added during data collection. We therefore consider the following equation for the time-evolution of the eigenvectors:

**Proposition 6.** *Let the eigenvalues of $\Sigma$ be ordered such that $\lambda_1 > \lambda_2 > \cdots > \lambda_n$. If $\lambda_1 \gg \lambda_j, \forall j \neq 1$, then the rate of rotation of the dominant eigenvector of $\Sigma$ is dominated by the corresponding entries of $Q^TJ^TQ$.*

*Proof.* Returning to the diagonalised Lyapunov equation we have (for $i \neq j$)

$$(M\Lambda - \Lambda M)_{ij} + \left(\frac{\mathrm{d}\Lambda}{\mathrm{d}t}\right)_{ij} = (Q^TJQ\Lambda + \Lambda Q^TJ^TQ + Q^TBQ)_{ij} \tag{S20}$$

$$(M\Lambda - \Lambda M)_{ij} = (Q^TJQ\Lambda + \Lambda Q^TJ^TQ + Q^TBQ)_{ij} \tag{S21}$$

$$\implies \left(Q^T\frac{\mathrm{d}Q}{\mathrm{d}t}\right)_{ij} = \frac{\lambda_j}{\lambda_j - \lambda_i}(Q^TJQ)_{ij} + \frac{\lambda_i}{\lambda_j - \lambda_i}(Q^TJ^TQ)_{ij} + \frac{1}{\lambda_j - \lambda_i}(Q^TBQ)_{ij} \tag{S22}$$

Assuming that $\lambda_1 \gg \lambda_j, \forall j \neq 1$, as seen in other applied studies in the literature[1,2], we consider the case that $i = 1, j > 1$. Then, the prefactors $\frac{\lambda_j}{\lambda_j - \lambda_1} \to -1, \frac{1}{\lambda_j - \lambda_1} \to -\frac{1}{\lambda_1}$ asymptotically, and the derivative simplifies to:

$$\left(Q^T\frac{\mathrm{d}Q}{\mathrm{d}t}\right)_{1j} \approx -(Q^TJ^TQ)_{1j} - \frac{1}{\lambda_1}(Q^TBQ)_{1j} \tag{S23}$$

$$\approx -(Q^TJ^TQ)_{1j} + O\left(\frac{1}{\lambda_1}\right). \tag{S24}$$

☐

This is similar to the expressions given in[1,3] for the evolution of the basis in the case of a stationary distribution/constant $J$ and $B$. Intuitively, this coincides with most of the variation in the system happening along one eigendirection, and could possibly relate to the system exhibiting oscillatory behaviour around a manifold (as expected from standard bifurcation theory). Similarly, it is possible to derive an evolution equation for the eigenvalues under the same assumption:

**Proposition 7.** *If $\lambda_1 \gg \lambda_j, \forall j \neq 1$, then $\frac{\mathrm{d}\lambda_1}{\mathrm{d}t}$ is driven primarily by the corresponding symmetric entry of $J$ when projected onto $Q$.*

*Proof.* Returning again to the differential equation for $\Sigma$ but now considering the diagonal entries, we have

$$(M\Lambda - \Lambda M)_{ii} + \left(\frac{\mathrm{d}\Lambda}{\mathrm{d}t}\right)_{ii} = (Q^T JQ\Lambda + \Lambda Q^T J^T Q + Q^T BQ)_{ii} \tag{S25}$$

$$\left(\frac{\mathrm{d}\Lambda}{\mathrm{d}t}\right)_{ii} = (Q^T JQ)_{ii}\lambda_i + \lambda_i(Q^T J^T Q)_{ii} + (Q^T BQ)_{ii} \tag{S26}$$

$$\implies \frac{\mathrm{d}\lambda_1}{\mathrm{d}t} = \lambda_1(Q^T JQ + Q^T J^T Q)_{11} + (Q^T BQ)_{11} \tag{S27}$$

$$= 2\lambda_1(Q^T JQ)_{11} + (Q^T BQ)_{11}. \tag{S28}$$

□
